# Disentangling the effect of measures, variants and vaccines on SARS-CoV-2 Infections in England: A dynamic intensity model

**DOI:** 10.1101/2022.03.09.22272165

**Authors:** Otilia Boldea, Adriana Cornea-Madeira, Joao Madeira

## Abstract

In this paper, we estimate the path of daily SARS-CoV-2 infections in England from the beginning of the pandemic until the end of 2021. We employ a dynamic intensity model, where the mean intensity conditional on the past depends both on past intensity of infections and past realised infections. The model parameters are time-varying and we employ a multiplicative specification along with logistic transition functions to disentangle the time-varying effects of non-pharmaceutical policy interventions, of different variants and of protection (waning) of vaccines/boosters. We show that earlier interventions and vaccinations are key to containing an infection wave. We consider several scenarios that account for more infectious variants and different protection levels of vaccines/boosters. These scenarios show that, as vaccine protection wanes, containing a new wave in infections and an associated increase in hospitalisations in the near future will require further booster campaigns and/or non-pharmaceutical interventions.

## 1 Introduction

In this paper, we use data on SARS-CoV-2 infections in England to estimate a time series model where the intensity of infections depends on both the level and intensity of past infections. We use this model to quantify the impact of the Omicron BA.1/BA.2 sub-variants and of the waning of immunity from vaccines/boosters on the COVID-19 epidemic in England, and to assess the timing and intensity of non-pharmaceutical interventions (NPIs) and further booster campaigns that may still be needed in 2022 to curb future infection waves. We additionally quantify the hospitalisation waves associated with new infection waves to show that further infections waves still require interventions.

There are two main challenges when fitting a model of COVID-19 to the data. First, the true number of cases is not observed and the ratio of unreported to reported cases varies over time, due to both changes in testing capacity and in testing behavior. Some econometric studies ignore unreported cases and model only reported cases (Jiang et al., 2020, Liu et al., 2021, Khismatullina and Vogt, 2021, Lee et al., 2021); this can lead to inconsistent parameter estimates or to serious mid and long-term forecasting errors, depending on the goal of the study (Korolev, 2021). Other studies employ various strategies to identify the share of unreported cases: Li et al. (2020) and Hortaçsu et al. (2021) identify the unreported cases through their mobility across regions; Arias et al. (2021), Rozhnova et al. (2021), Viana et al. (2021) and Toulis (2021) use random sample serology tests; Gourieroux and Jasiak (2020) use parameteric time-varying transition probabilities, and Sonabend et al. (2021) use random tests in the population. We use the last identification strategy, as England runs a bi-weekly random sample population survey based on polymerase chain reaction (PCR) tests, from which we construct a time-varying ratio of total to reported cases and apply it to (delayed) daily reported cases to approximate the total daily cases.

The second main challenge is model complexity. Most of the large-scale stochastic epidemiological modelling papers that address the effectiveness of policy interventions compartmentalise the population into susceptible, exposed, infected, recovered and possibly other states such as hospitalisations or deaths. These models are necessarily complex over longer periods of time, because, for example, only modelling infections in vaccinated or waned vaccinated typically require introducing another set of compartments for each and therefore more unobservables (see, e.g., Sonabend et al., 2021, and the citations therein). Because data on each infection type is typically not available at higher frequency, several parameters in these models are unidentified and require calibration. With these additional calibrations, these models can be estimated by Bayesian filtering methods, although, due to nonlinearity compound with several (unobserved) state variables and many parameters, their estimation can pose substantial computational challenges.^1^

To reduce model complexity while allowing for vaccination and its waning, we propose a different approach, where the population is not compartmentalised, and the effect of seasonality, vaccination and waning enters the model parameters multiplicatively to the effect of variants of concern and that of non-pharmaceutical interventions. To that end, we employ a dynamic intensity model with time-varying parameters, where infections are assumed to follow a negative binomial distribution to allow for overdispersion due to superspreader events.^2^ This model is akin to integer generalized autoregressive conditional heteroskedasticity (INGARCH) models but instead of modelling variance clustering, it models intensity clustering: when the intensity of the infection process is high, it stays high for a while and it is reinforced by the level of past infections.^3^

The model parameters vary based on individuals’ behavior as a result of NPIs. We estimate both the timing and the magnitude of the behavioral response following NPIs in a similar fashion to Rozhnova et al. (2021) and Viana et al. (2021). The parameters also vary with vaccination, and we estimate the intensity reduction from vaccination based on the vaccine schedule and the total infections. This allows us to combine different administered vaccines into a single vaccine intensity reduction parameter without requiring separate data on infections of vaccinated and non-vaccinated individuals, data which is not available at daily frequency. The parameters also vary with variants of concern and we estimate the timing and the effect of these variants in a similar fashion to Viana et al. (2021). The seasonality cannot be identified separately and is calibrated based on previous studies.

The advantage of our model over more complex models is that it can be estimated relatively quickly, and therefore can be used in real-time to inform policy makers on the interventions needed and their timing, depending on new variants and (waning) effects of boosters. The disadvantage compared to more complex epidemiology models is that it cannot explicitly account for the share of the susceptible population entering and changing during an infection wave. However, since a large fraction of individuals are susceptible to Omicron BA.1 and BA.2 sub-variants, regardless of their vaccination or previous infection status, our model provides a good approximation to the path of infections in the near future.

As the infection data is not stationary over long periods, we estimate the model via Bayesian Hamiltonian Monte Carlo methods. Disentangling the effect of vaccines and boosters from those of variants and NPIs allows us to employ counterfactuals and provide scenarios for the future six months, both using NPIs and further booster campaigns.

Our counterfactuals shows that the timing of NPIs and of vaccines and boosters is key in curbing infections waves. We find that the recent Omicron wave could have been substantially mitigated by earlier timing and faster speed of vaccine and booster schedules or two weeks of lockdown in mid-December 2021. Our scenarios show that another wave can happen in the coming months due to booster waning, and its occurrence depends on a range of factors. First, on the transmissibility of the Omicron BA.2 sub-variant: if its intensity increase relative to Omicron BA.1 is large, a new wave can occur as early as March 2022. Second, on the choice of NPIs: maintaining semi-lockdown restrictions from mid-December of 2021 may delay the next infection wave to the summer. Third, on the effectiveness of boosters: if the booster intensity reduction is sufficiently high, under some scenarios another infection wave is substantially delayed.

We then examine the implications these scenarios have for new hospital admissions. Our projected hospital admissions track well observed hospital admissions, and we show that new hospitalisations rise steeply, shortly after the start of another infection wave.

The rest of the paper is organised as follows. Section 2 describes the model. Section 3.1 describes the data. Section 3.2 contains estimation results. Section 3.3 presents the counterfactual analysis, and Section 3.4 provides projections of daily infections for the spring and summer of 2022. In Section 3.5 we approximate the daily new hospitalisations as a results of infections for counterfactuals and projection scenario. Section 4 concludes. The Supplementary Appendix provides plots of parameter posterior distributions along with parameter identification results obtained by simulation, as well as additional counterfactuals and scenarios.

## 2 Model

The model for daily total COVID-19 cases (reported and unreported), *y*_*t*_, is a negative binomial conditional response model:

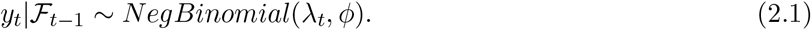

The probability distribution function is given by

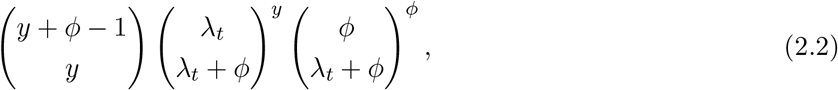

with *λ*_*t*_ ∈ ℝ^+^, *ϕ* ∈ ℝ^+^, *y* ∈ ℕ. The mean and the variance are given by *E*[*y*_*t*_|ℱ_*t*−1_] = *λ*_*t*_ and 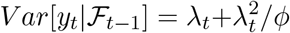, where 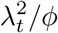 is the additional variance above the mean *λ*_*t*_, ℱ_*t*−1_ = {*y*_*t*−1_, *λ*_*t*−1_, *y*_*t*−2_, *λ*_*t*−2_,.. and

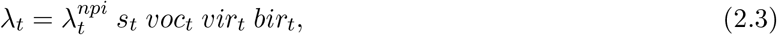

where 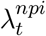 is the daily intensity of infections due to NPIs (either restrictions or relaxation of restrictions), *s*_*t*_ is seasonality, and *voc*_*t*_, *vir*_*t*_ and *bir*_*t*_ are changes in intensity due to variants of concern, vaccines and boosters.

To account for the seasonal pattern of SARS-CoV-2 (by which transmission is lower in summer and higher in winter), we define the sinusoidal function *s*_*t*_ (Liu et al., 2021):

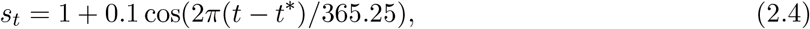

where *t*^*^ is January 1 (due to coldest weather).

To account for the increase in intensity due to variants of concern, we define:

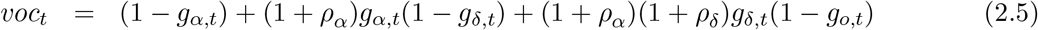

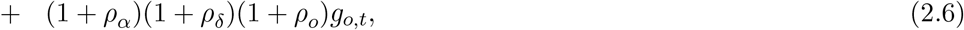

where the parameters *ρ*_*α*_, *ρ*_*δ*_ and *ρ*_*o*_ represent the relative intensity increase of the Alpha, Delta and Omicron BA.1 variants that became dominant in England in January 2021, June 2021 and December 2021 respectively. The intensity increase as the new variants take over is described using the logistic functions:

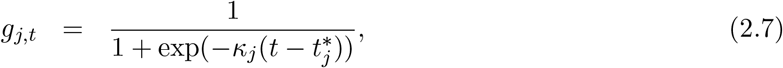

where *j* = *α, δ, o* are the variants of concern, *κ*_*j*_ is the steepness of the logistic function and 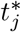 is the midpoint of the logistic function. The functions *g*_*j,t*_ can be interpreted as probabilities of contracting the new variant, which increase over time, while *ρ*_*j*_ can be interpreted as the relative intensity increase when the new variant completely takes over. Therefore, as described in Section 3.1, we fitted the logistic functions (2.7) to external gene sequencing data as in Viana et al. (2021) and Hansen (2021), while *ρ*_*j*_ is estimated directly from fitting infection data.^4^

The effect of vaccinations and boosters and their waning is modelled as:

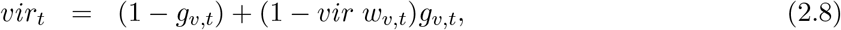

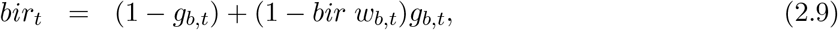

where *vir*_*t*_ and *bir*_*t*_ describe the vaccine/booster-induced intensity reduction. If there are no vaccinated individuals, *vir*_*t*_ and *bir*_*t*_ are equal to 1. As more vaccines are administered, *vir*_*t*_ and *bir*_*t*_ decrease to (1 − *vir w*_*v,t*_)*g*_*v,t*_ and (1 − *bir w*_*b,t*_)*g*_*b,t*_ respectively, where *vir* is the vaccine (two doses) intensity reduction parameter and *bir* is the booster intensity reduction parameter. The transition from no vaccination to vaccination is described by the logistic functions *g*_*v,t*_ and *g*_*b,t*_:

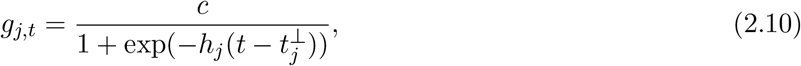

with *j* = *v, b*, where *h*_*j*_ is the steepness, and 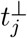 is the midpoint of the transition function. We assume *c* = 0.7 (the fraction of the total population of England that had the 2nd dose of the vaccine by the beginning of January 2022 when our sample ends). The logistic transition function for the vaccine and booster uptake *g*_*v,t*_ and *g*_*b,t*_ are fitted to total share of daily vaccinations and boosters administered, as explained in Section 3.1. Following Keeling et al. (2021), we introduce waning of vaccine protection against infection through the exponential function:

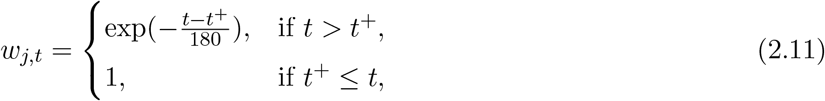

where *j* = *v, b*. For the estimation, we assume that the waning of vaccines starts on June 28, 2021, hence *t*^+^ = June 28, 2021 (6 months after the first 2nd dose vaccine was administered December 29, 2020). For the booster, we do not assume waning in the estimation since our estimation ends in December 24, 2021 (3 months after the first dose of the booster was administered in September 16, 2021). However, in the counterfactuals (Section 3.3), we assume that the boosters wane after four, five and six months, while in the scenarios (Section 3.4), we assume that boosters wane after five months, and the results for six months are relegated to the Supplementary Appendix.

We specify 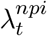 as:

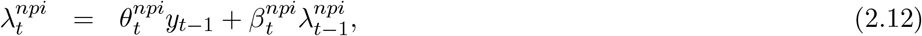

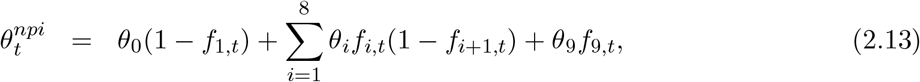

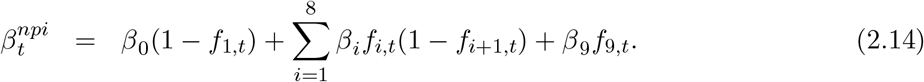

As can be seen from (2.12), 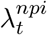 is triggered by the previous day infections (*y*_*t*−1_) and previous day intensity 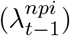. The parameters, *θ*_*i*_ ≥ 0 and *β*_*i*_ ≥ 0, *i* = 0, …, 9, associated with *y*_*t*−1_ and 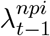 change in each regime by *γ*_*i*_ and *ω*_*i*_ respectively: *θ*_*i*_ = *θ*_*i*−1_ +(−1)^*i*^*γ*_*i*_, *β*_*i*_ = *β*_*i*−1_ +(−1)^*i*^*ω*_*i*_ *i* = 1, …, 9, through logistic transition functions

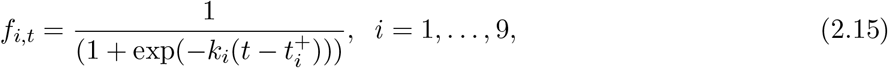

where the *k*_*i*_ describe the speed at which restrictions or relaxation measures are taken up by individuals, and 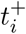 describe the mid-time of the take-up of a restriction/relaxation. The correspondence between each regime and NPIs is described in Table 1, where only the last regime does not refer to a NPI, but to a transition to school holidays; nevertheless, we refer to it for simplicity as an NPI regime. The *f*_*i,t*_ logistic functions are directly estimated within the model, and approximate the timing and the effect of individuals’ behavior following NPIs in a similar fashion to Rozhnova et al. (2021) and Viana et al. (2021). This feature is not present in any other studies that employ the INGARCH model, but is important because individuals might react ahead of measures, or might take time to adjust to measures.

**Table 1:** NPIs transition functions

| NPI transitions | Meaning |
| --- | --- |
| $f_{1,t}$ | first lockdown in 2020 to relaxation measures in the summer 2020 |
| $f_{2,t}$ | relaxation measures in the summer 2020 to second lockdown in November 2020 |
| $f_{3,t}$ | second lockdown to some relaxation measures before Christmas 2021 |
| $f_{4,t}$ | relaxation measures before Christmas 2020 to third lockdown in January 2021 |
| $f_{5,t}$ | lockdown in January 2021 to relaxation measures in Spring 2021 |
| $f_{6,t}$ | further relaxation measures and big gatherings<br>(the Euro 2020 football tournament end of June - beginning of July 2021) |
| $f_{7,t}$ | no big crowded events (July 2021, after the end of the Euro 2020 tournament) |
| $f_{8,t}$ | transition to a period with full relaxation (no restrictions) including<br>school/universities opening (from July 2021 to October 2021) |
| $f_{9,t}$ | transition from school opening to school holiday (after late October 2021) |

## 3 Results

### 3.1 Data and Estimation Method

#### Data on variants of concern

We obtained the weekly percentage of COVID-19 positive cases by gene pattern and Cycle threshold (Ct) value from the Office for National Statistics (ONS) Coronavirus (COVID-19) Infection Survey from England and we linearly interpolated them to obtain the daily percentage of COVID-19 positive cases. For the Alpha variant we used gene sequencing data from December 3, 2020 until January 10, 2021. The Alpha variant was identified if the ORF1ab and N genes were present. For the Delta variant we used gene sequencing data between April 26, 2021 and December 6, 2021. The Delta variant was identified if ORF1ab, N and S genes were present. For the Omicron BA.1 variant we used gene sequencing data between November 29, 2021 and January 3, 2022. The Omicron BA.1 variant was identified because of the absence of the S-gene in combination with the presence of the ORF1ab and N genes. The logistic functions (2.7) were fitted to the daily percentage of COVID-19 positive cases by gene.

#### Data on vaccines and boosters

We used daily observations for England from the official COVID-19 in the UK dashboard on new people vaccinated with the second dose and new people receiving a booster dose (from December 29, 2021 until January 13, 2022, and from September 16, 2021 until January 13, 2022 respectively). The logistic functions (2.10) were fitted to the daily cumulative number of people vaccinated with the second dose and receiving the booster.

#### Constructed Daily Cases

For the model given in (2.3), our data is from May 3, 2020 until January 22, 2022, a total of *N* = 630 daily infections.^5^ The estimation of (2.3) is carried until December 24, 2021 and the data from December 25, 2021 until January 22, 2022 is used to assess the out of sample fit of the model’s projections (in Section 3.4). The daily infections *y*_*t*_ in (2.12) refer to the reported and unreported cases, 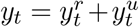. For the reported daily infections 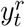 we used new cases by specimen date obtained from the official Coronavirus (COVID-19) in the UK dashboard. To approximate the total daily infections, we proceeded as follows. Denote by 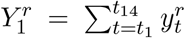 the reported cases by specimen date for the period May 3-16, 2020, …, 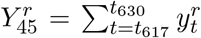 the reported cases by specimen date for the period January 9-22, 2022. The reported cases are considered to be reported with a delay of 2 days since the onset of the symptoms (Casey-Bryars et al., 2021). Denote by 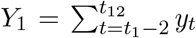 the total infections for the period May 1 - 14, 2021, …, 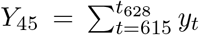 the total infections for the period January 7 - 20, 2022. From the ONS Coronavirus (COVID-19) Infection Survey we have the estimated percent (say *p*_*j*_) of the population that had COVID-19 for a time period of 14 days. Then, *Y*_*j*_ = *p*_*j*_ × 56, 550, 138*/*100, where 56, 550, 138 is the population in England (based on the ONS mid-year population estimates, June 2020). We calculate 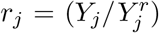, the ratio of total infections and reported infections in the two-week period *j, j* = 1, …, 45. To calculate the daily total cases we assume the daily ratio of total to reported infections within a 14-day period is equal to the two-week ratio corresponding to that 14-day period. Let 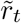 denote the daily ratio of total to reported cases; then the total cases are 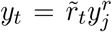. Note that we constructed daily data because some of the NPIs transition functions are too short to use biweekly data, and could not have been estimated otherwise.

The total and reported new cases are shown in Figure 1 below. The divergence between the two series is highest in times of high incidence, possibly due to limits to testing capacity, but also possibly due to testing behavior, suggesting that correcting for unreported cases is essential to remove the time-varying sample selection bias in reported cases.^6^

**Figure 1:**
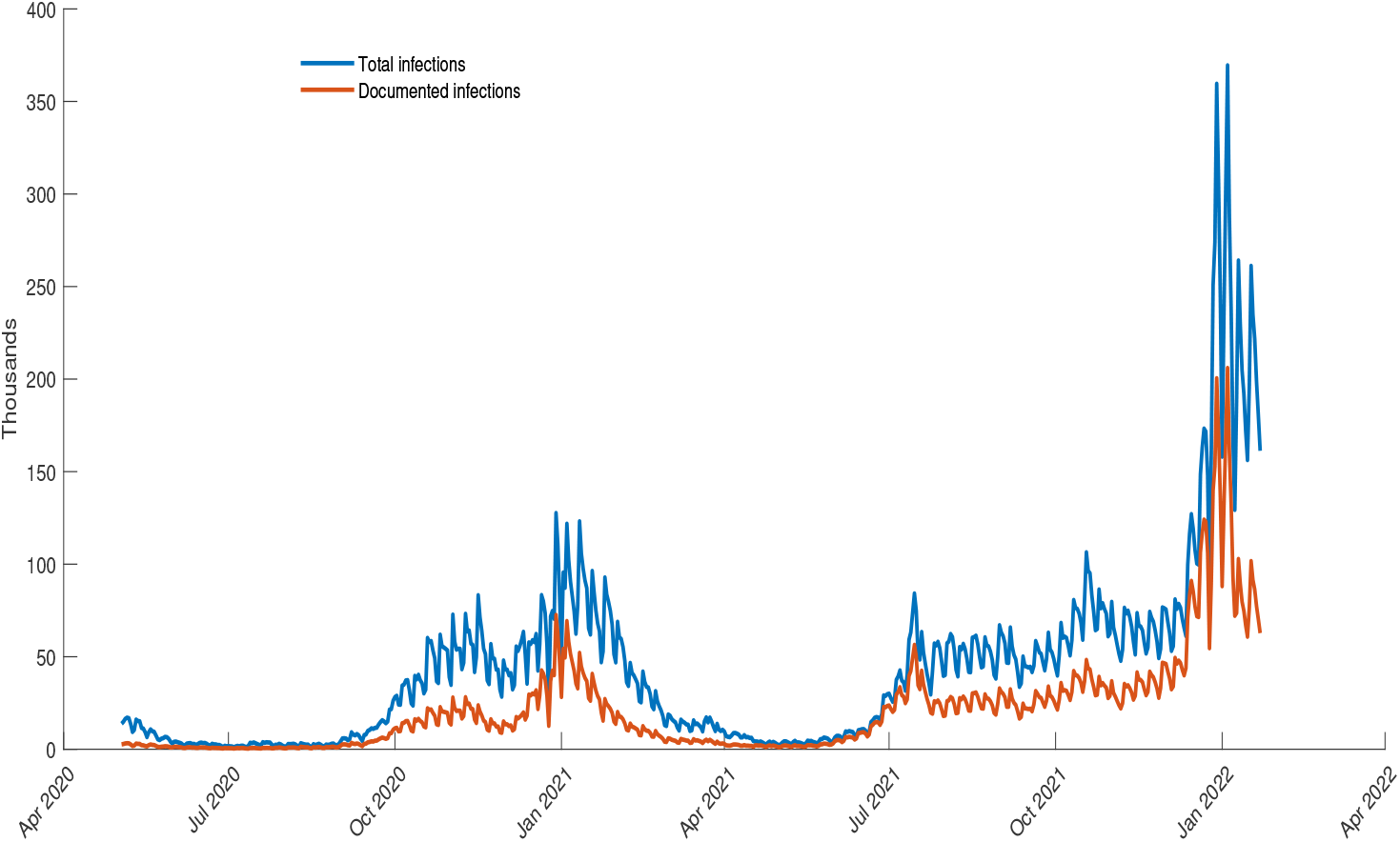
Total infections and reported infections between May 3, 2020 until January 22, 2022 in England

#### Estimation

Because total cases are not stationary, we use Bayesian estimation with Hamiltonian Monte Carlo (HMC) methods, implemented in R Stan, and for simplicity we also did so for stationary data such as the share of new variants or cumulative vaccine/boosters uptake. Neal (2011) and Fernández-Villaverde and Guerrón-Quintana (2021) provide a description of the HMC and highlight its computationally efficiency relative to traditional Markov Chain Monte Carlo (MCMC) methods, due to exploiting information from the gradient of the posterior, which reduces the correlation between successive parameter values in the Markov chain and therefore ensures that the Markov chain converges much faster than in MCMC.

Denoting by *x*_*j,t*_ the daily percentage of COVID-19 positive cases due to the new variant or the daily cumulated vaccine/booster uptake, we used 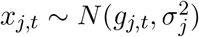, where *g*_*j,t*_ is given by (2.7) (with *j* = *α, δ, o*) or (2.10) (with *j* = *v, b*). Table 2 below lists the priors for all parameters - whether they are estimated using sequenced gene data, vaccine data, or infections - and motivates their choice.

**Table 2:** Prior distributions of the parameters in the model

| Parameter | Prior | Description |
| --- | --- | --- |
| $\theta_0$ | LogN(0, 1) | Vague prior (infections data) |
| $\gamma_i$ | LogN(0, 1) | $i = 1, \dots, 9$, vague prior (infections data) |
| $\beta_0$ | LogN(0, 1) | Vague prior (infections data) |
| $\omega_i$ | LogN(0, 1) | $i = 1, \dots, 9$, vague prior (infections data) |
| $\rho_\alpha$ | LogN(0, 0.5) | Vague prior for Alpha (infections data) |
| $\rho_\delta$ | LogN(0, 1) | Larger scale than for Alpha (infections data) |
| $\rho_o$ | LogN(2, 2) | Larger scale than for Delta (infections data) |
| $vir$ | Beta(5, 2) | Gives more probability mass to values larger than 0.5 (infections data) |
| $bir$ | Beta(5, 2) | Gives more probability mass to values larger than 0.5 (infections data) |
| $k_i$ | Exp(1) | $i = 1, \dots, 9$ (infections data) |
| $t_1^+$ | N(35,7) | 35 =June 6, 2020, relaxation in the summer 2020 (infections data) |
| $t_2^+$ | N(140,7) | 140 =September 19, 2020, transition to the second lockdown in November 2020 (infections data) |
| $t_3^+$ | N(200,7) | 200 =November 18, 2020, transition from the lockdown in November 2020 to relaxation (infections data) |
| $t_4^+$ | N(230,7) | 230 =December 18, 2020, transition from relaxation to lockdown in January 2021 (infections data) |
| $t_5^+$ | N(270,7) | 270 =January 27, 2021, transition from the lockdown in January 2021 to relaxation (infections data) |
| $t_6^+$ | N(425,7) | 425 =July 1, 2021, transition to large crowds events (Euro 2021) (infections data) |
| $t_7^+$ | N(440,7) | 440 =July 17, 2021 transition to no large crowd events (infections data) |
| $t_8^+$ | N(515,7) | 515 =September 29, 2021 transition to a period with full relaxation and schools/universities opening (infections data) |
| $t_9^+$ | N(555,7) | 555 =November 8, 2021, transition from school opening to school holiday (infections data) |
| $\phi$ | LogN(0,1) | Vague prior (infections data) |
| $\kappa_i$ | Exp(1) | As in Rozhnova et al. (2021), $\kappa_i = 1$ means lift-up $\sim 6$ days ($i = \alpha, \delta, o$) (gene data) |
| $t_\alpha^*$ | N(10,7) | 10 =December 12, 2020, around the date when Alpha became dominant (gene data) |
| $t_\delta^*$ | N(10,7) | 10 =April 26, 2021, around the date the Delta emerged (gene data) |
| $t_o^*$ | N(17,7) | 17 =December 15, 2021, around the date Omicron became dominant (gene data) |
| $\sigma_j$ | LogN(0,1) | Vague prior, $j = \alpha, \delta, o, v, b$ (gene/vaccination/booster data) |
| $h_i$ | Exp(1) | $i = v, b$ (vaccination/booster data) |
| $t_v^\perp$ | N(190,7) | 190 =July 6, 2021, around the date when 50% of the population had the vaccine (2nd dose) (vaccination data) |
| $t_b^\perp$ | N(106,7) | 106 =December 12, 2021, around the date 50% of the population had the booster (booster data) |

### 3.2 Posterior estimates

The transition functions for sequenced gene data and vaccination are shown in Figures 2a and 2b, and the fitted total infections in Figure 3. The posterior medians along with their 90% credible intervals are listed in Table 3. The estimated steepness of the transition functions based on sequenced gene data is the highest for the Omicron BA.1 variant, 0.1508, while the steepness of transition function for the Delta variant, 0.0377, is slightly higher than for the Alpha variant which is 0.0372. The estimated transition functions for the vaccines and boosters from the vaccination data show that the uptake of the booster is faster than the uptake of the vaccine 2nd dose.

**Table 3:**
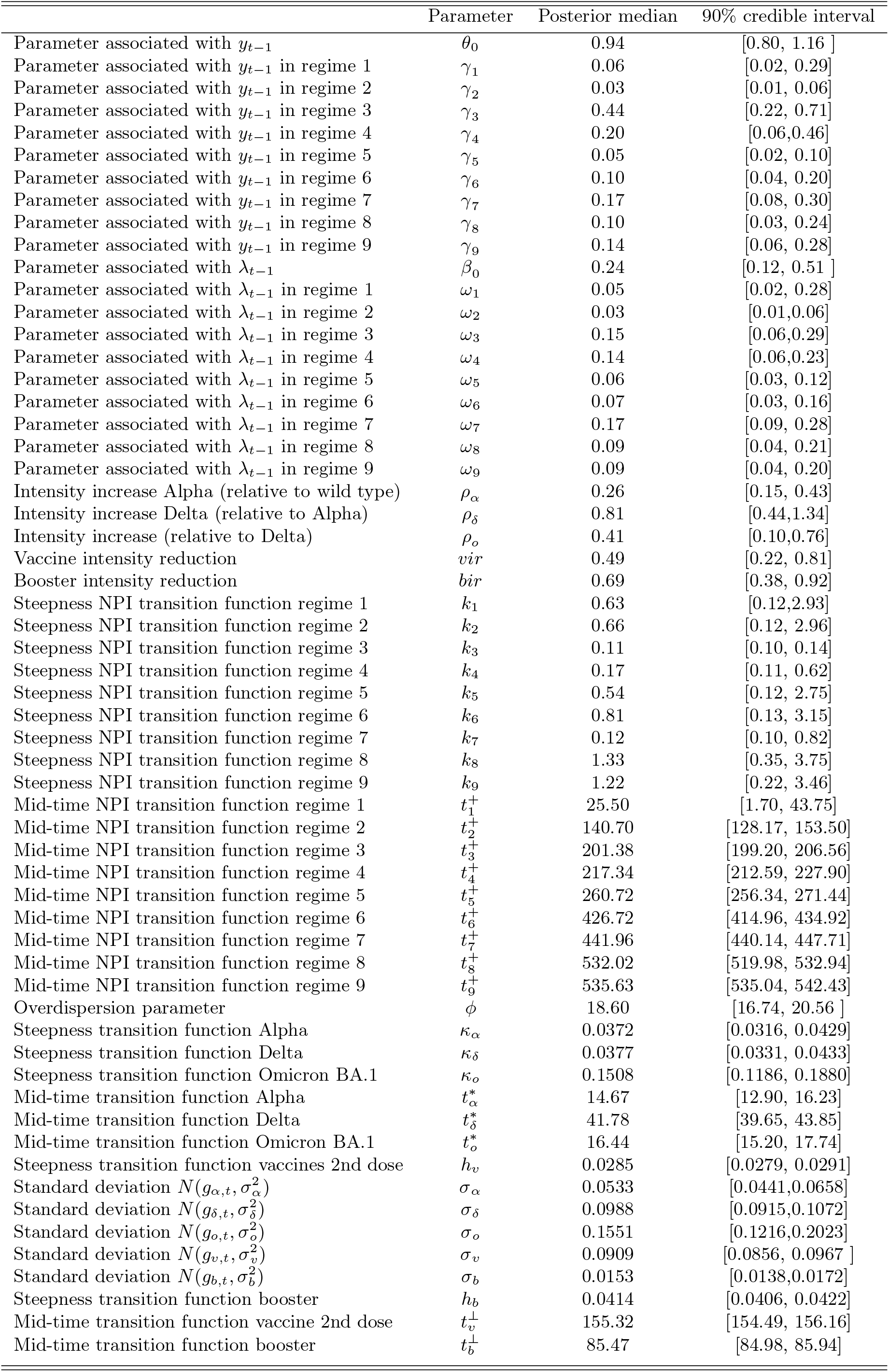
Posterior medians

**Figure 2.**
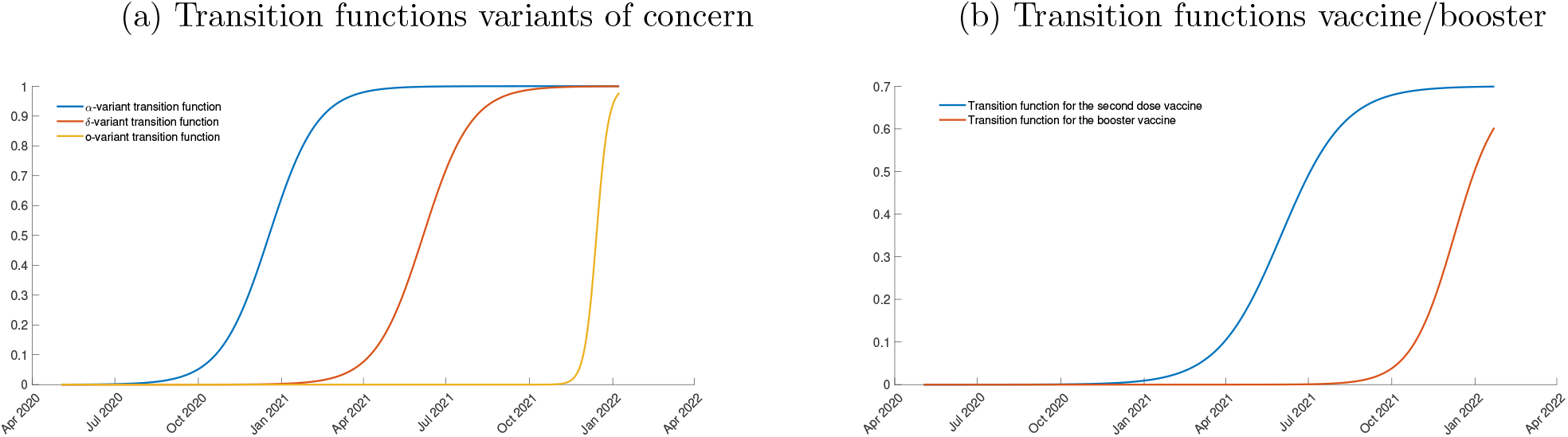

**Figure 3:**
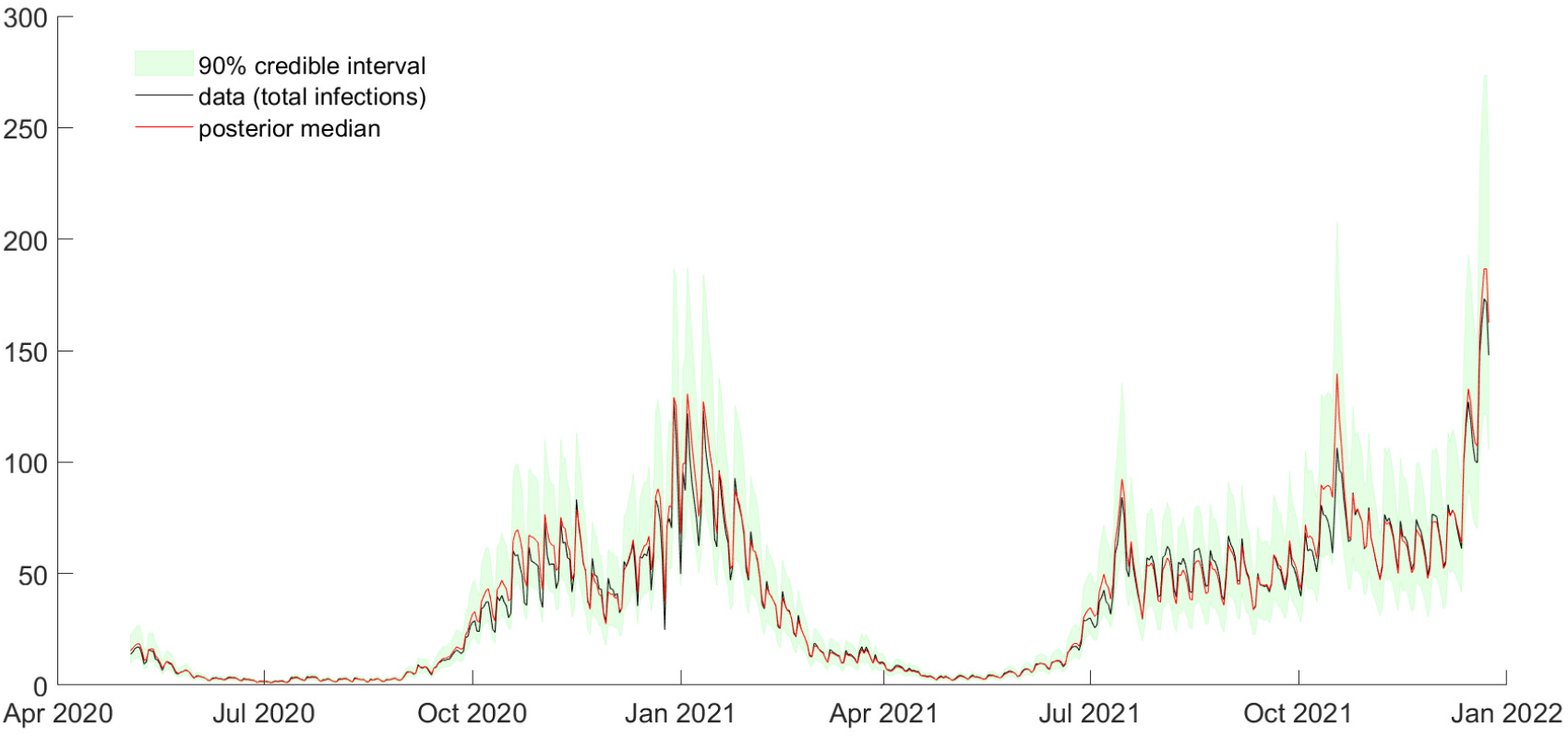
Fitted versus total cases

The rest of the parameters are estimated within the dynamic intensity model. As can be seen from Table 3 above, the Alpha, Delta and Omicron BA.1 variants result in 26%, 81% and 41% higher relative intensity. The vaccine intensity reduction parameter estimates *vir* and *bir* are 49% and 69% respectively.^7^ The posterior medians for steepness of the transition functions in the regimes 8 and 9 (since the full relaxation in the summer 2021) are the highest (1.33 and 1.22 respectively). The overdispersion parameter estimate is large, showing that a model without overdispersion would fit the data poorly.

The way the model is written may suggest that some parameters only enter multiplicatively and cannot be identified; however, the regimes over which these parameters are identified only partially overlap, and this is ensured by gluing the transition functions, allowing identification from the time variation in the non-overlap periods. In the Supplementary Appendix, Section S2, we show that the posteriors for most parameters are tighter than their priors, and further demonstrate identification through simulations.

The model implies that 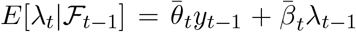, where 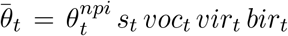 and 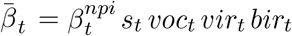. In Figures 4 and 5 below, we show the estimated time evolution of the median posterior estimates of 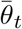 and 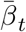 plotted against the variants, vaccinations and the timing of various measures. In these figures Steps 1 - 4 refer to the steps in the roadmap out of the third lockdown (that took place in early 2021) in England. We further plot the contribution to the estimated 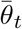 of the estimates of 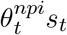 and 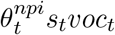, and similarly for 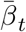. Figure 4 shows that in principle, NPIs were effective should they have been implemented against the wild-type variant, but because of new variants, their effectiveness dropped over time. The figure also shows that vaccines and boosters substantially mitigated this drop. We see a drop in 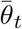 in the summer of 2020, followed by an increase in autumn 2020. This second wave is brought under control by a second lockdown, but 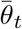 starts to increase again as the Alpha variant takes over. At the same time, vaccinations begin and this tempers down the increase until the Delta variant takes over and large events such as the Euro-2020 cup are allowed, in which period the transmission soars. With these events no longer in place, the transmission decreases again but then schools open, and we see another steep surge, which is tempered by school holidays, but most importantly by boosters being widely administered. We see a similar evolution for 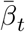 in Figure 5, except that as the Omicron variant becomes dominant, this parameter stays low. This can be explained by the fact that 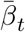 measures the dependence of infections on the recent past infections rather than the level of infections yesterday. This dependence becomes less important with Omicron, as this variant is widely shown to generate immune escape, so that infections in the recent past, with a previous variant, play a less important role than they did at providing protection against infection compared to the case when other variants were dominant. Nevertheless, the estimate of 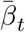 is not close to zero, indicating that this dependence is not negligible. This also motivates our use of the reinforcing term 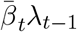: without it, the dependence on infections on the recent past infections cannot be easily quantified.

**Figure 4:**
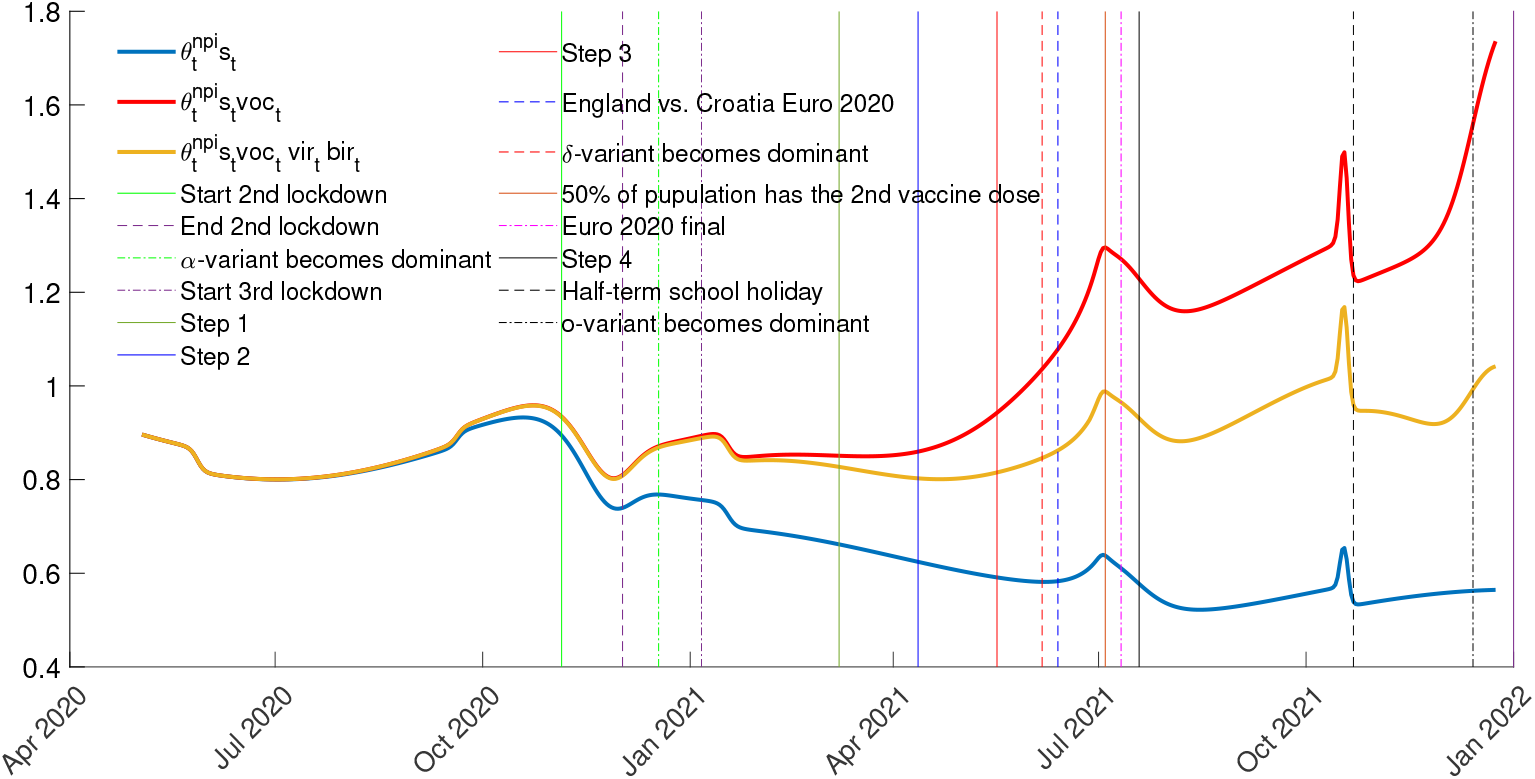
Estimated time evolution of posterior median estimates of the parameters associated with previous day infections *y*_*t*−1_

**Figure 5:**
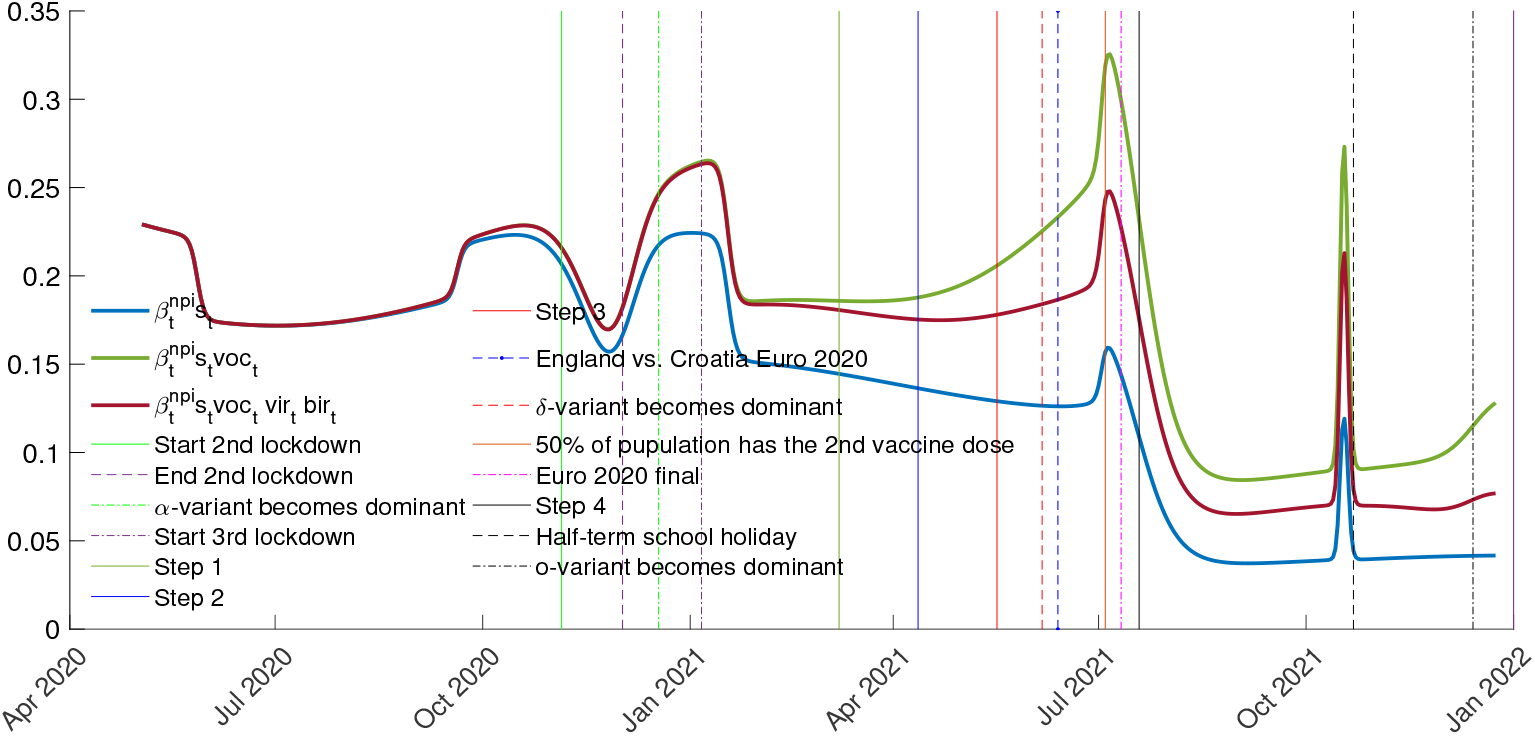
Estimated time evolution of posterior median estimates of the parameters associated with previous day intensity *λ*_*t*−1_

### 3.3 Counterfactual analysis

We use the estimates from Section 3.2 to run counterfactuals regarding the timing and intensity of booster campaigns (Figures 6 and 7) and NPIs (Figures 8 and 9). In all figures, the shaded areas represent the interquartile range from 4000 negative binomial draws (in the Supplementary Appendix, Section S3, we included the same figures, but with the lower 5% to the upper 95% quantiles). The projected daily median infections (solid lines) is given by the median from these 4000 draws.

**Figure 6:**
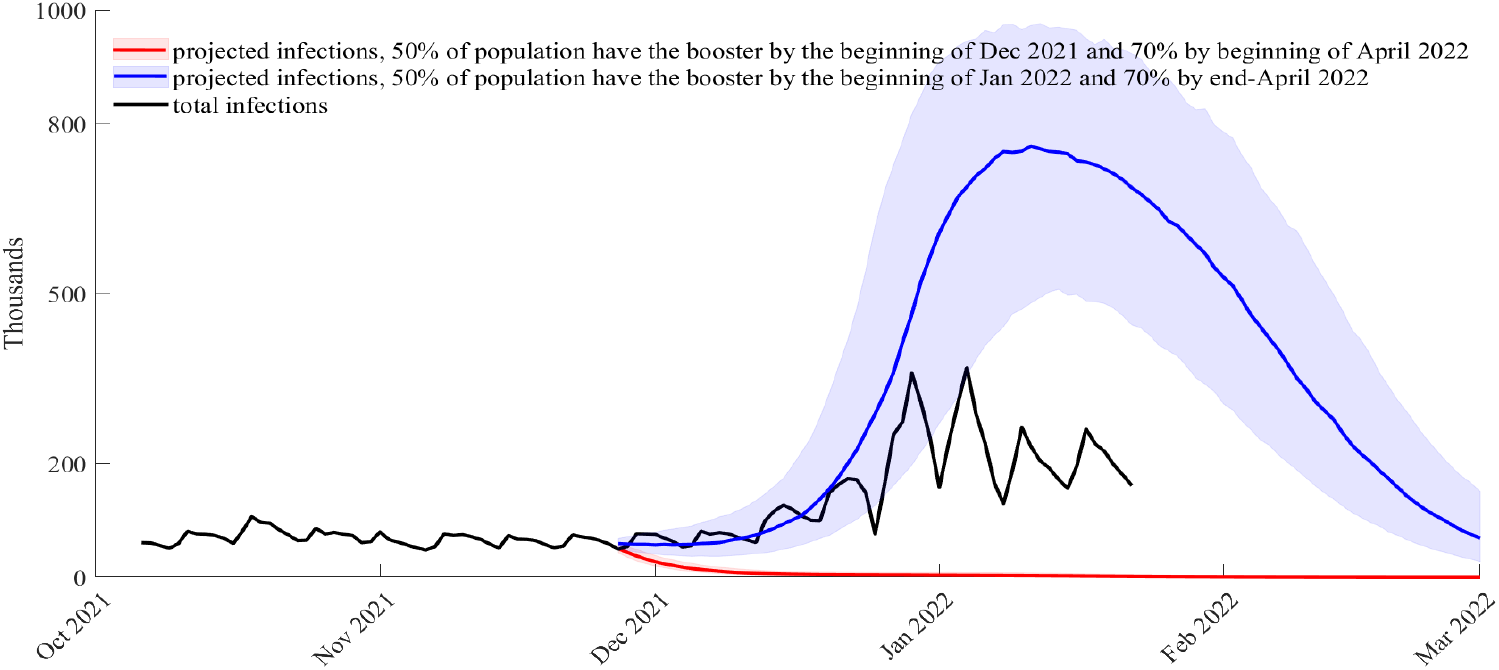
Counterfactual when the vaccine booster campaign starts on August 16, 2021 and the population is reached faster (red) or slower (blue); projection of daily infections from November 27, 2021; *bir* = 0.69 (posterior median) and *ρ*_*o*_ = 0.41 (posterior median)

**Figure 7:**
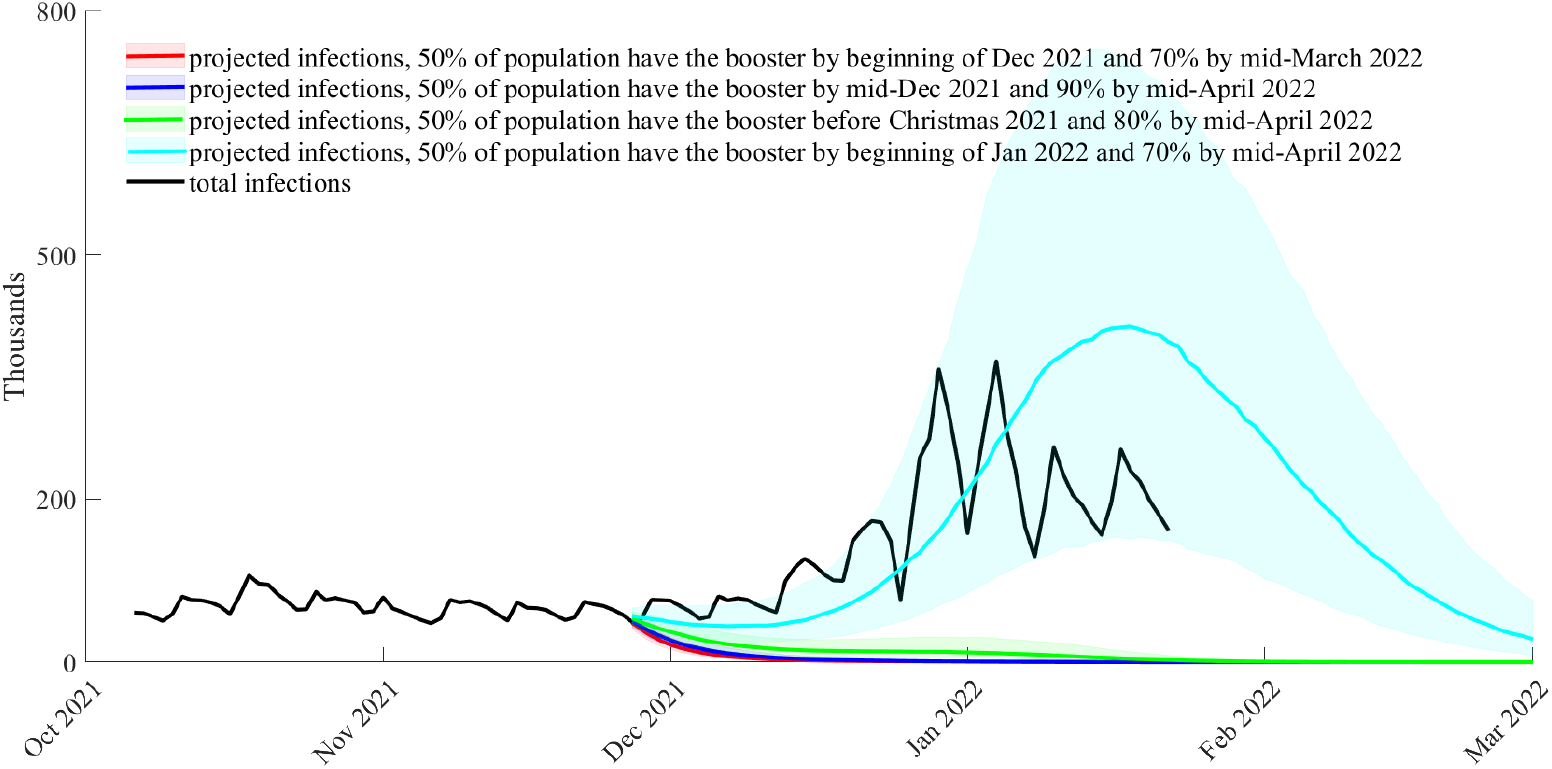
Counterfactual when the vaccine booster campaign starts on September 16, 2021 and population is reached at different speeds; projection of daily infections from November 27, 2021; *bir* = 0.69 (posterior median) and *ρ*_*o*_ = 0.41 (posterior median)

**Figure 8:**
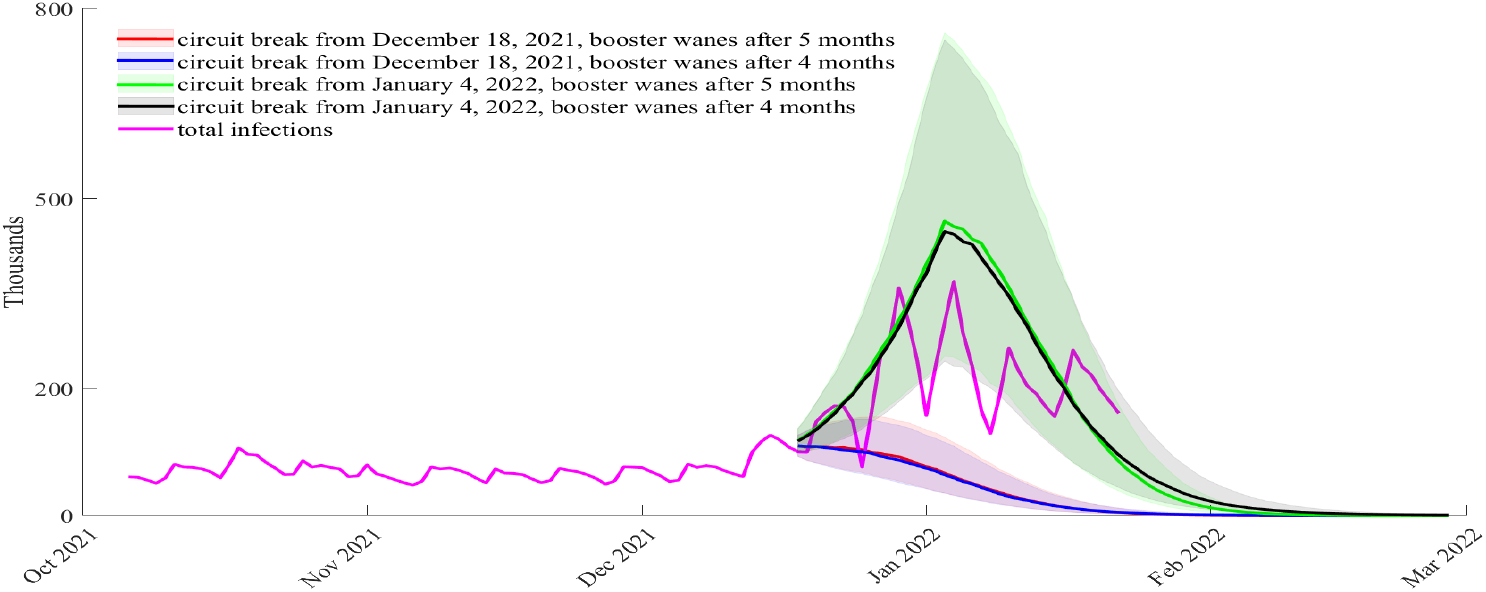
Counterfactual when there is a circuit break (2 weeks hard lockdown) from December 18, 2021 or January 4, 2022 (peak of infections); projection of daily infections from December 18, 2021; *bir* = 0.69 (posterior median) and *ρ*_*o*_ = 0.41 (posterior median)

**Figure 9:**
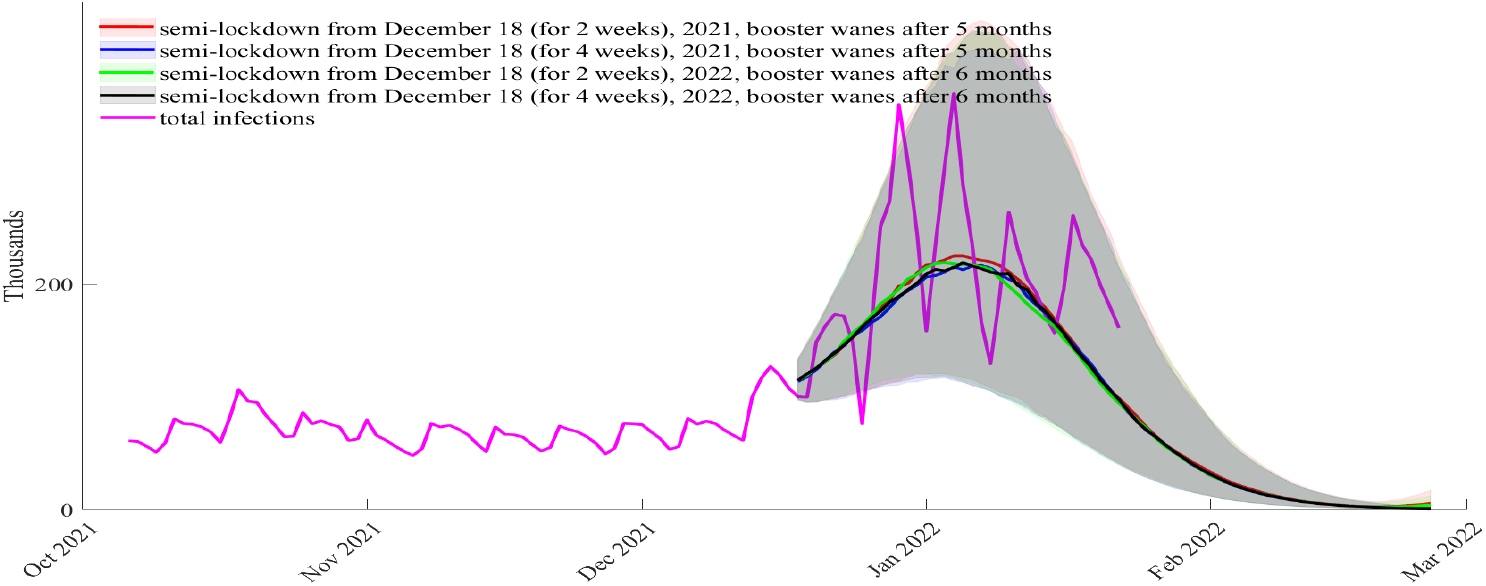
Counterfactual when there is a semi-lockdown from December 18, 2021; projection of daily infections from December 18, 2021; *bir* = 0.69 (posterior median) and *ρ*_*o*_ = 0.41 (posterior median)

Denote by 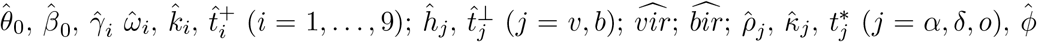 the posterior medians from Table 3, which are used to obtain the estimates 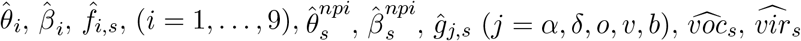 and 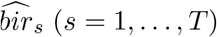, the in-sample time evolution of the parameters. Then, for *t* = *T* + *𝓁* (*𝓁* ≥ 1), a draw from the negative binomial is:

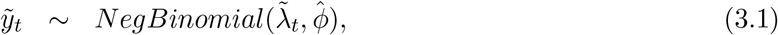

and the parameters’ time evolution for this draw are given by

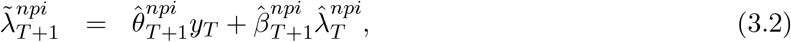

for *𝓁* = 1, and by

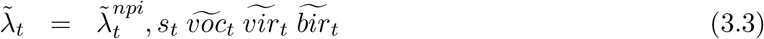

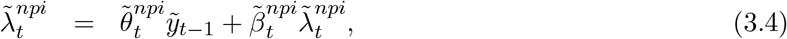

for *𝓁* ≥ 2.

Figure 6 below shows what would have happened if the booster campaign had started in mid-August 2021 rather than mid-September 2021. In this counterfactual, *t* = *T* + 1 corresponds to November 27, 2021 (when the first case of Omicron BA.1 variant was identified in England), and for *t* = *T* + 1, …, we obtain the out-of-sample parameters evolution: 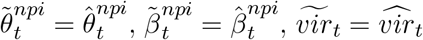,

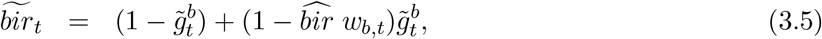

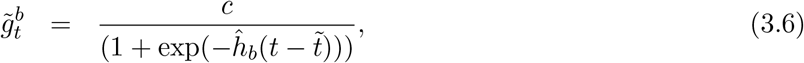

where 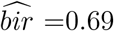, *ĥ*_*b*_ = 0.0414 (posterior medians for *bir* and *h*_*b*_ from Table 3), and *c* = 0.7 (70% of the population is assumed to be reached by the vaccine/booster campaigns, as we considered in the estimation of the model in Section 3.2). The booster waning in (3.5), *w*_*b,t*_, starts 5 months after August 16, 2021. It is calculated as in (2.10) with *t*^+^ = January 15, 2021. For the mid-time in the transition function of the booster, (3.6), we consider the case when 50% of the population is reached relatively fast, that is, in 85 days, exactly like the current booster campaign that started in mid-September 2021 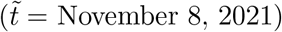. We also consider the case when 50% of the population is reached later than 85 days since the start of the booster campaign in mid-August, more exactly in 120 days 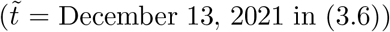. In Figure 6 we present the projected daily infections from November 27, 2021 (when the first case of Omicron BA.1 variant was identified in England). The black line shows the total daily infections. The red line shows the posterior median prediction for the scenario in which 50% of the population received the booster by the start of December 2021. The blue line shows the posterior median prediction for the scenario in which 50% of the population received the booster by the start of January 2021. Hence, this figure shows that had the booster campaign started one month earlier and reached quickly a significant fraction of the population, the winter infection wave could have been avoided. However, an early start would not have been sufficient to avoid a winter wave if the booster uptake was not fast enough.

Figure 7 below shows a hypothetical scenario in which the booster campaign started as it actually occurred, but the population is reached at different speeds and/or more people received the booster. In this counterfactual, *t* = *T* + 1 in (3.1) corresponds to November 27, 2021 (when the first case of Omicron BA.1 variant was identified in England). For *t* = *T* + 1, … we obtain the out-of-sample parameters evolution as follows: 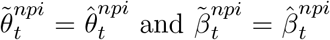. As one cannot have a booster without being fully vaccinated first, for vaccines, we use the transition function estimated from the vaccination data, but with total vaccinated population share *c* ∈ {0.7, 0.8, 0.9}, where *c* = 0.7 is what actually occurred, and *c >* 0.7 are hypothetical scenarios.^8^ More exactly, in (3.3) we use:

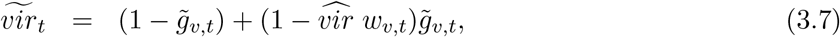

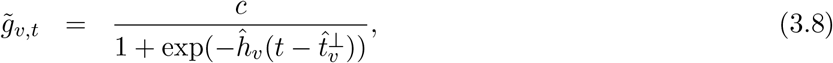

where the 2nd dose vaccine waning, *w*_*v,t*_, starts on June 28, 2021 (as considered in the estimation, Table 3), and *c* = 0.7, 0.8, 0.9. For the booster, we use (3.5) and (3.6), but with 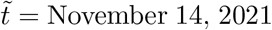 and *c* = 0.7 (70 % of the population is reached, red line from Figure 7). This scenario corresponds to the situation when 50% of the population is reached by the beginning of December 2021. We also consider the hypothetical scenario when 50% of the population is reached later in December 2021 and by the beginning of January 2022, in which case we take 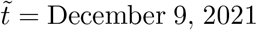 (for the blue, green and light blue lines from Figure 7), with different speeds: *c* = 0.9 (90% of the population is reached, blue line), *c* = 0.8 (80% of the population is reached, green line), *c* = 0.7 (70% of the population is reached, light blue). In Figure 7 we present the projected daily infections from November 27, 2021 (when the first case of Omicron BA.1 variant was identified in England). Figure 7 confirms that the speed of the booster campaigns would have been key to maintain the spread of Omicron. The estimated model predicts that had 50% of the population received a booster before Christmas 2021 then the winter wave driven by the spread of Omicron could have been avoided. If 50% of the population is boosted by early January (which is what occurred) then the estimated model predicts a winter wave similar to what was observed up to the start of 2022 with a peak being reached in mid-January. In reality the number of infections peaked in early January which suggests that the measures adopted during December 2021 (after mid-December masks became mandatory in most public indoor venues, individuals were advised to work from home and proof of vaccination was required to enter nightclubs or attend large gatherings) to contain the spread of Omicron did have an impact (note that the projected daily median infections in the scenarios considered start in November 27, 2021 which is prior to the adoption of restriction measures in December 2021).

In Figures 8 and 9 below we present respectively a counterfactual analysis with a circuit breaker (two weeks of hard-lockdown as recommended by one member of the Scientific Advisory Group for Emergencies in England) and a semi-lockdown (similar to what was implemented by the government in England after mid-December of 2021). In Figures 8 and 9 the projected daily infections are from December 18, 2021 (*t* = *T* +1 in (3.1) corresponds to this date). In particular, 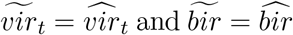 with booster waning function *w*_*v,t*_ calculated as in (2.11) with *t*^+^ = January 15, 2021 (waning starts 4 months after mid-September 2021), *t*^+^ = February 14, 2021 (waning starts 5 months after mid-September 2021) and *t*^+^ = March 16, 2021 (waning starts 6 months after mid-September 2021). We consider the following hypothetical evolution of the parameters due to the NPIs:

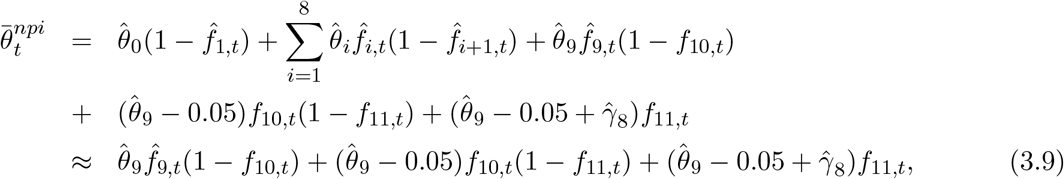

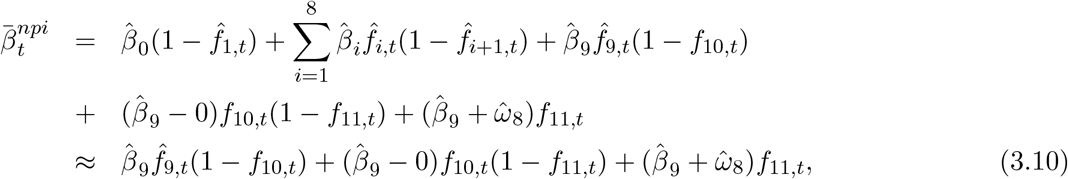

where the results in (3.9) and (3.10) follow from the fact that for *t* = *T* + 1, …, the transition function 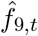 is the dominant one and the transition functions for the previous regimes have no impact. In (3.9) and (3.10) above, *f*_10,*t*_ is the transition function from relaxation to hard lockdown or semi-lockdown:

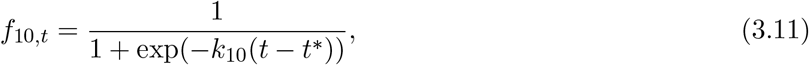

with *t*^*^ =December 22, 2021 (for the circuit breaker starting on December 18, 2021), *t*^*^ =January 8, 2022 (for the circuit breaker starting on January 4, 2022 when total infections reach their peak), and *t*^*^ = December 28, 2021 (for the semi-lockdown that starts on December 18, 2021). The steepness of the transition function (3.17) is considered *k*_10_ = 0.1 (similar to 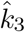 the estimate of steepness of the transition function 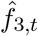 from relaxation to hard-lockdown in November 2020 in Table 3). For the hard lockdown, the midpoint of the transition function is reached 4 days after the lockdown is imposed, while for the semi-lockdown the midpoint is reached after 10 days. Hence, the transition function is steeper for the hard lockdown compared to the semi-lockdown. The exit from lockdown in period with relaxation is captured though the transition function

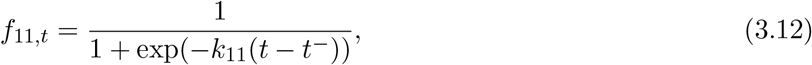

with steepness equal to *k*_11_ = 1 (≈ 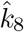 the steepness of the transition function 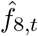 to a period of full relaxation in the summer and Autumn 2021, see Table 3). Moreover, *t*^−^ = February 20, 2022 (for the circuit breaker in December 18 with exit in 2 weeks), *t*^−^ = March 9, 2022 (for the circuit breaker in January 4 with exit in 2 weeks), *t*^−^ = March 10, 2022 (for the semi-lockdown in December 18 with an exit in 2 weeks), *t*^−^ = March 23, 2022 (for the semi-lockdown in December 18 with an exit of 4 weeks). For all scenarios in Figures S11 and S12 we assume that the exit is in 2 or 4 weeks after the lockdown is imposed, but we allow for the fact that restrictions are usually lifted gradually, not in one go. To account for this, the midpoint of the transition function from lockdown to relaxation is reached 50 days after the measures are lifted. The choice of 50 days is motivated by the fact that between the midpoint of 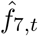 and that of 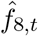 (between the big sport event at the beginning of July 2021 and the peak of infections in late October 2021 when full relaxation measures were in place and the Delta variant was dominant) there are 90 days. The results from Figures S11 and S12 are similar if the midpoint of *f*_11,*t*_ is reached after 100 days (if restrictions are lifted slower). As seen from (3.9), the parameter associated with previous day infections, *y*_*t*−1_, transitions from 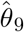 to 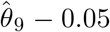 during lockdown (to reflect that the lockdown reduces the infections). The choice of 0.05 is similar to 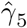 (see 3) in regime 5 (after the third lockdown that started in January 2021). As seen from (3.16) the parameter associated with past daily intensity *λ*_*t*−1_ remains unchanged. This is motivated by the fact that starting with the period when Delta became dominant, and in particular when the Omicron BA.1 became dominant, the importance of *λ*_*t*−1_ has diminished while the importance of *y*_*t*−1_ in triggering new infections has increased, reflecting the fact that Omicron is more transmissible than previous variants. Once the lockdown ends, the parameters increase by 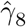 and 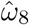 (the posterior medians of the parameters associated with the NPI in the summer 2021 when full relaxation measures were in place).

Figures 8 and 9 also demonstrate that the timing of adoption of restriction measures is key. If a circuit breaker lockdown had been implemented in mid-December 2021 then the estimated model predicts that the increase in cases in early winter due to Omicron could have been avoided. However, a late circuit breaker lockdown or a semi-lockdown of either 2 or 4 weeks would not have prevented the winter wave. Note that the semi-lockdown scenario of 4 weeks is close to what the government implemented (the government restrictions were put in place between mid-December to late January, approximately 6 weeks) and the estimated model projections track well the total realised infections. The similarity between the counterfactual cases of 2 and 4 weeks semi-lockdown indicates that perhaps the government could have ended restrictions earlier than it did and that would not have resulted in a significant increase in infections.

### 3.4 Scenarios

In this section, we provide scenarios for the evolution of total COVID-19 cases in the next six months. As in Section 3.3 the shaded areas in all figures represent the interquartile range from 4000 negative binomial draws (in the Appendix, Section S4, we included the same figures, but with the lower 5% to the upper 95% quantiles). The projected daily infections are given by the median from these 4000 draws. The draws are obtained as described at the beginning of Section 3.3 and are based on (3.1)-(3.4) with *t* = *T* + 1 corresponding to December 25, 2021. Moreover,

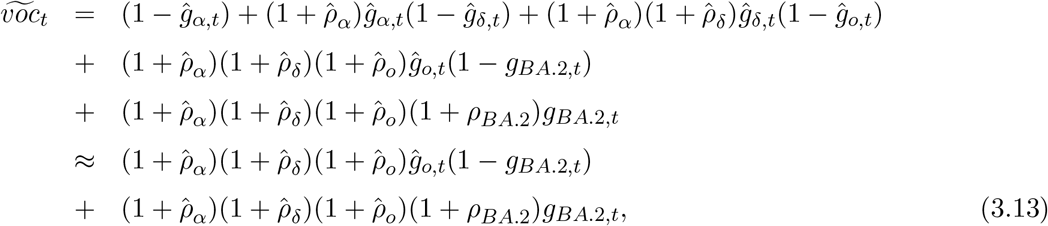

where the result in (3.13) follows from the fact that from the summer 2021 the Delta variant is dominant; *ρ*_*BA*.2_ is the intensity increase parameter for the Omicron BA.2 variant relative to the Omicron BA.1 variant, and *g*_*BA*.2,*t*_ is the transition function of the Omicron BA.2 variant

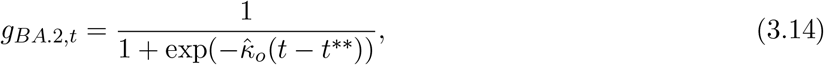

with 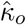 the steepness of the transition function for the Omicron BA.1 variant *ĝ*_*o,t*_, and the mid-time of *g*_*BA*.2,*t*_ is *t*^**^ = February 5, 2022, that is 12 days after the first cases of infections with the Omicron BA.2 variant were genomically confirmed in England. For the Omicron BA.1 variant the mid-time in the transition function was estimated to be 16 days after the first case appeared in England (November 27, 2021). Thus, we assume that the BA.2 variant spreads more rapidly than the BA.1 variant.

In obtaining the draws 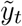, we also considered:

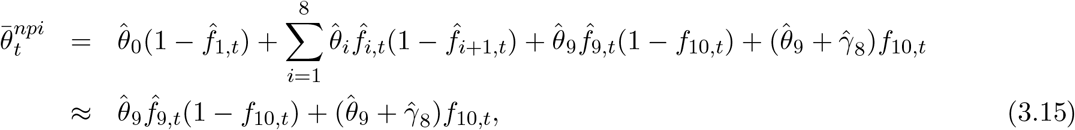

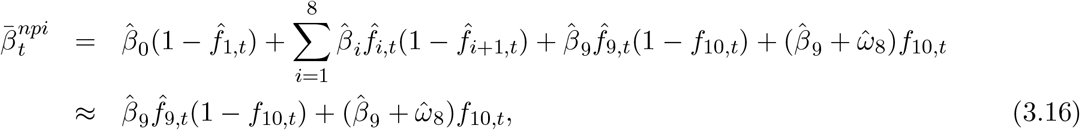

where *f*_10,*t*_ is the transition function from some relaxation measures to full relaxation

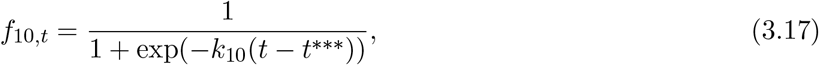

with *k*_10_ = 1 (≈ 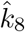 the steepness of the transition function 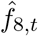 to a period of full relaxation in the summer and Autumn 2021, see Table 3). We consider two cases for *t*^***^. The first one corresponds to a scenario with an early impact of measures lifted: *t*^***^ = March 13, 2022 (50 days after January 27 2021 when measures of Plan B were lifted).^9^ The second case corresponds to a scenario with a late impact of measures lifted: *t*^***^ = May 6, 2022 (100 days after January 27 2021 when measures of Plan B were lifted).

Finally, to obtain the draws 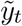 we have 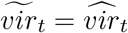, and

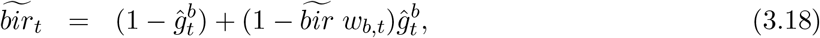

where 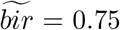 (Figures S13-S15 below) and 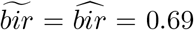 (posterior median) (Figures S17-S20 in Section S5 from the Supplementary Appendix). The higher *bir* is chosen because: (a) the scenarios with *bir* = 0.69 seem pessimistic when plotted against infections in February 2022, which were not used in the estimation, but just plotted out of sample; (b) it partially compensates for the fact that we do not account for temporary immunity in the model, but previously acquired infections with Omicron BA.1 may temporarily protect against infection with Omicron BA.2, as suggested by a recent Danish study - Lyngse et al.(2022). Therefore, the scenarios considered are a baseline case in which the BA.2 sub-variant does not become dominant (Figure 10), (*ρ*_*BA*.2_ = 0) and three cases where the BA.2 sub-variant leads to *ρ*_*BA*.2_ ∈ {5%, 10%, 20%} intensity increase compared Omicron BA.1 (Figures 11-13). We assume that the booster wanes in 5 months (Figures 10-13), and the Supplementary Appendix (Section S5, Figures S21-S23) shows the same scenarios but with waning after 6 months.

**Figure 10:**
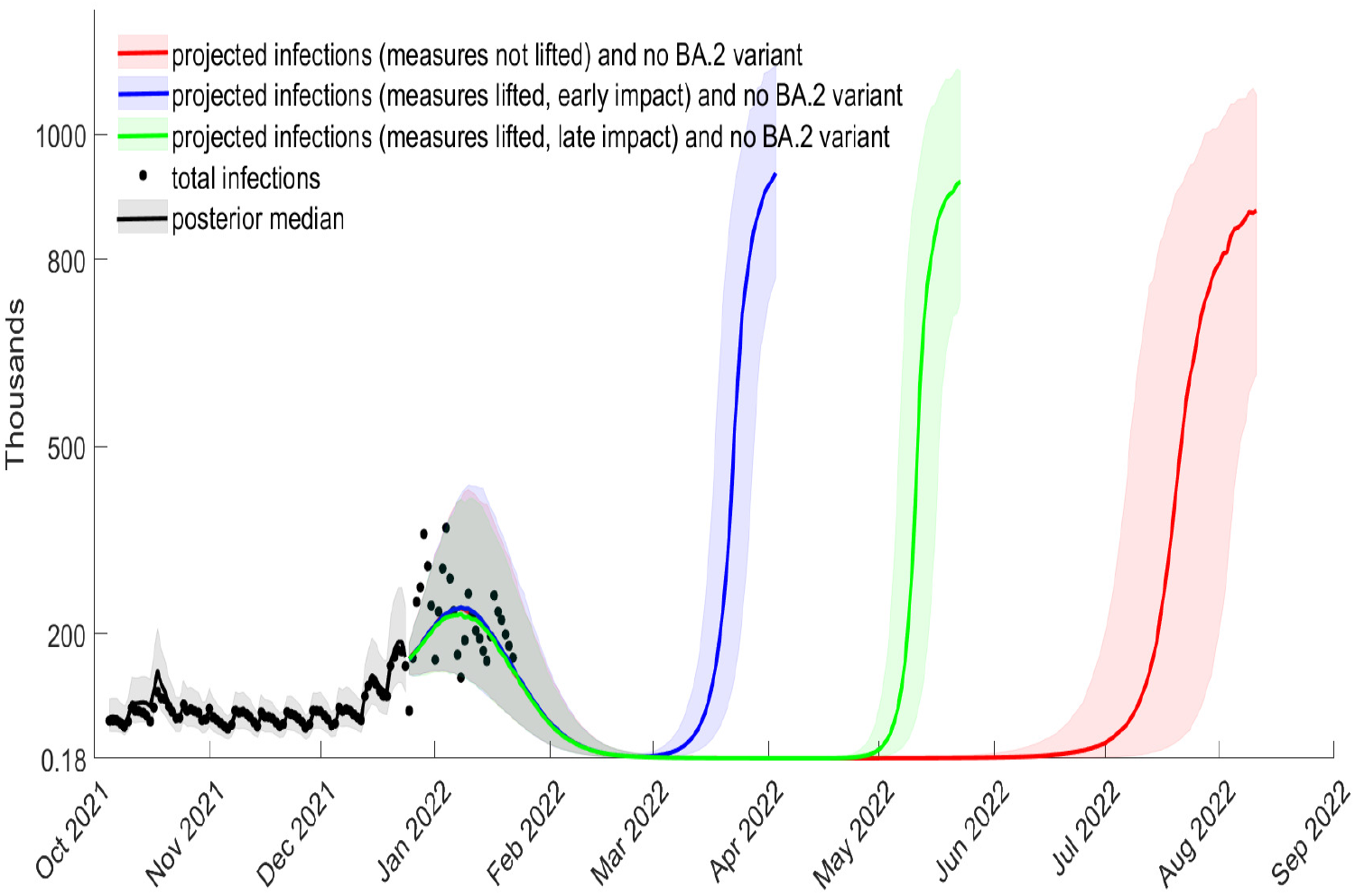
Projected total cases from December 25, 2021, waning of boosters after 5 months, *bir* = 0.75, *ρ*_*o*_ = 0.41 (posterior median), no increase in relative BA.2 intensity

**Figure 11:**
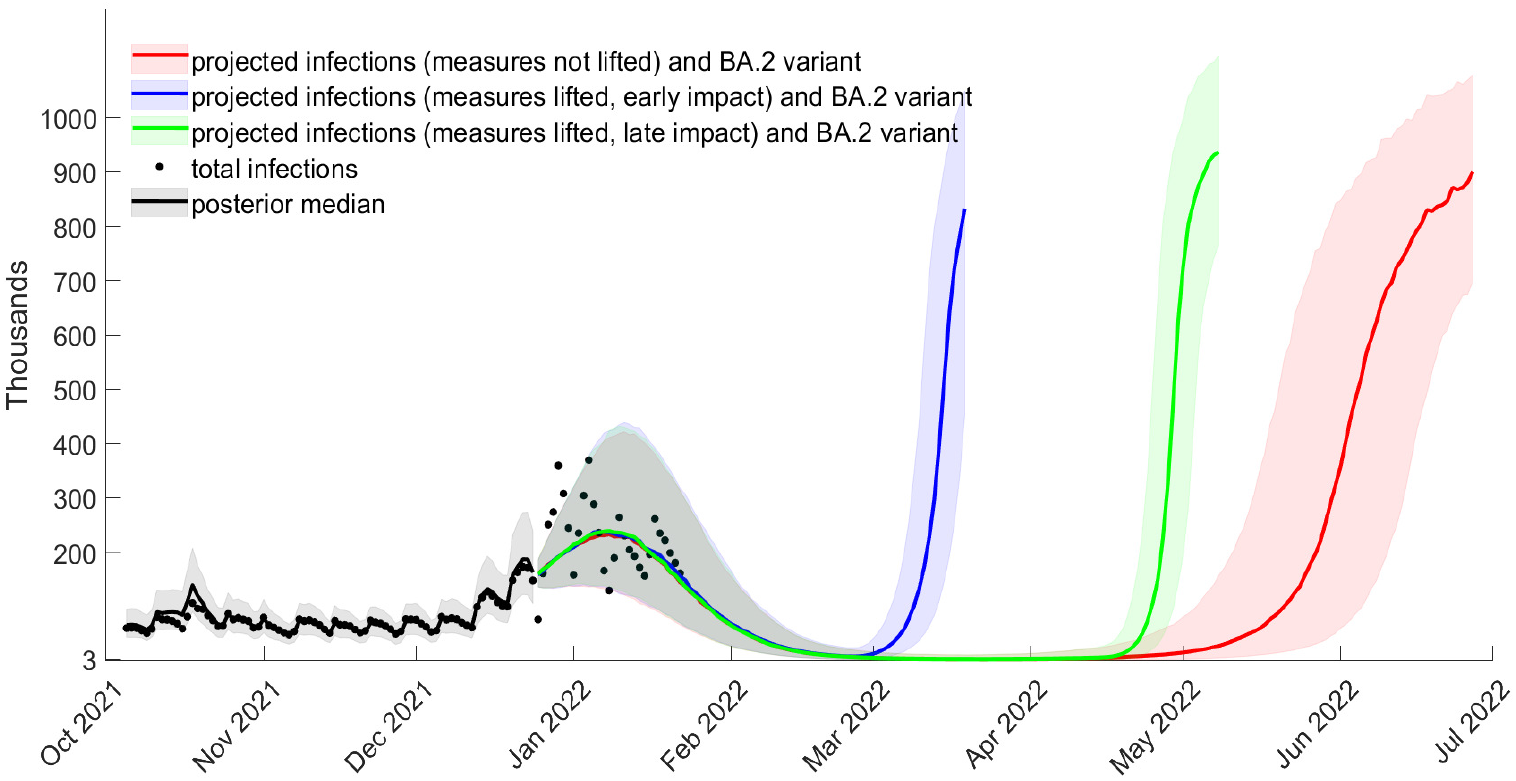
Projected total cases from December 25, 2021, waning of boosters after 5 months, *bir* = 0.75, *ρ*_*o*_ = 0.41 (posterior median), relative BA.2 intensity increase *ρ*_*BA*.2_ = 5%

In all scenarios we consider projections for whether NPIs are not lifted, lifted having an early impact on infections and lifted having a late impact on infections. The case in which the lifting of restrictions has early impact represents a scenario in which individuals quickly start taking advantage of the greater freedoms allowed and the case of a late impact represents a scenario in which individuals remain cautious. Black dots indicate the observed total infections and the black line the in-sample posterior median for the estimated model from Section 2.

Figure 10 shows that the estimated model projections match well the observed peak in infections observed in early January, 2022 with infections projected to continue falling until they reach very low levels by March. However, we can also observe that due to waning of boosters after 5 months, infections increase again. The timing for the emergence of a new wave will depend on restriction measures. Maintaining restrictions similar to those implemented from mid-December, 2021 to late January, 2022 could mean delaying the new wave to the summer. Lifting the measures can lead to a new wave emerging in either early spring (if there is an early impact from lifting restrictions) or late spring (if there is a late impact from lifting restrictions).

Figure 11 shows that if the Omicron BA.2 sub-variant has an intensity increase of *ρ*_*BA*.2_ = 5% relative to Omicron BA.1 (Figure 11), then the timing of the projected new wave changes relative to what was shown Figure 10: with late impact from lifting restrictions, a new wave starts in mid-April (rather than May) and in the case of no lifting of restrictions a new wave starts in late May, 2022 (rather than July). If *ρ*_*BA*.2_ = 10% (Figure 12) then for both the case of late impact from lifting restrictions and no lifting of restrictions a new wave starts in late mid-April. If *ρ*_*BA*.2_ = 20% (Figure 13) then a new wave is predicted to start in March (even if the restrictions imposed in mid-December of 2021 were to be kept).

**Figure 12:**
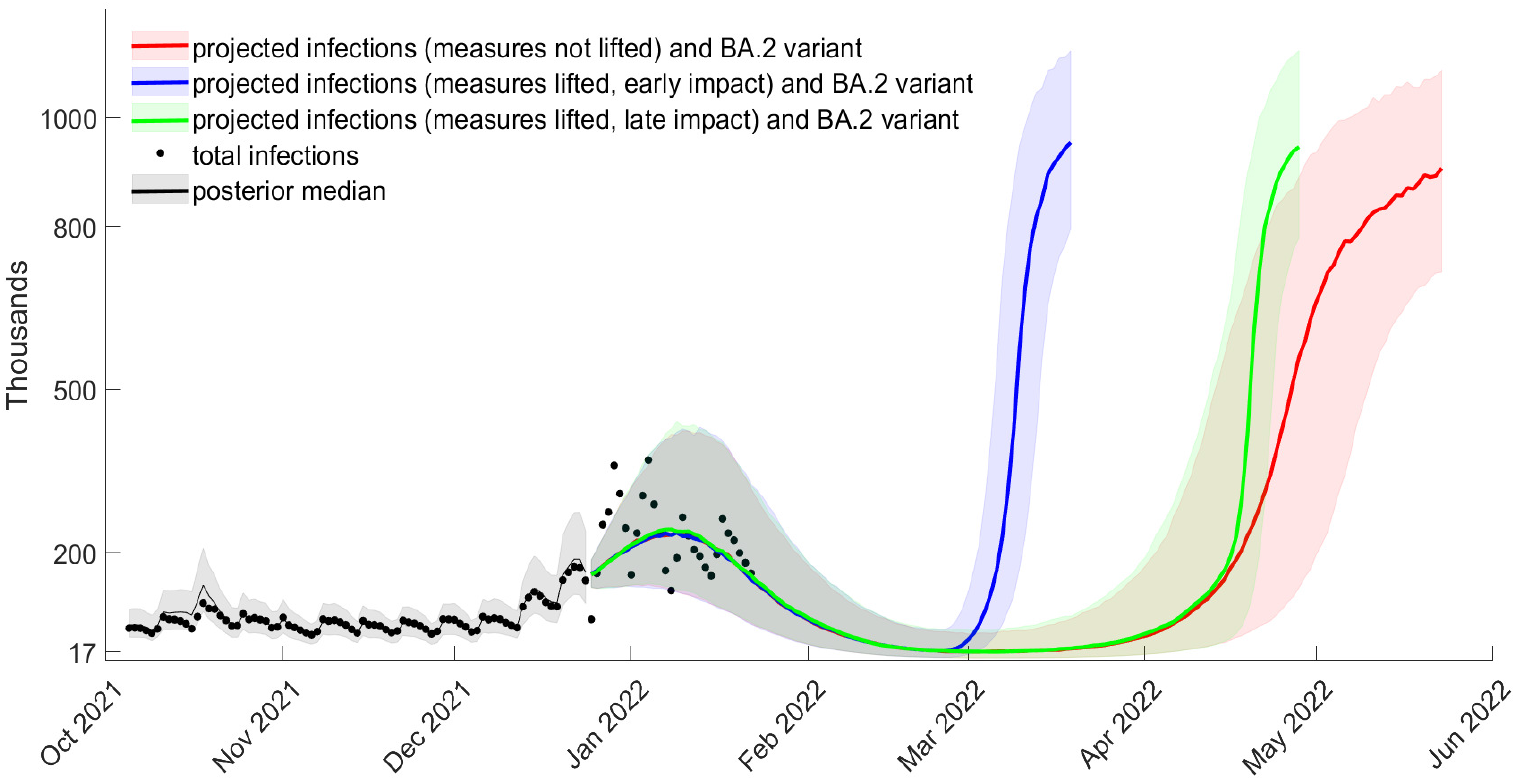
Projected total cases from December 25, 2021, waning of boosters after 5 months, *bir* = 0.75, *ρ*_*o*_ = 0.41 (posterior median), relative BA.2 intensity increase *ρ*_*BA*.2_ = 10%

**Figure 13:**
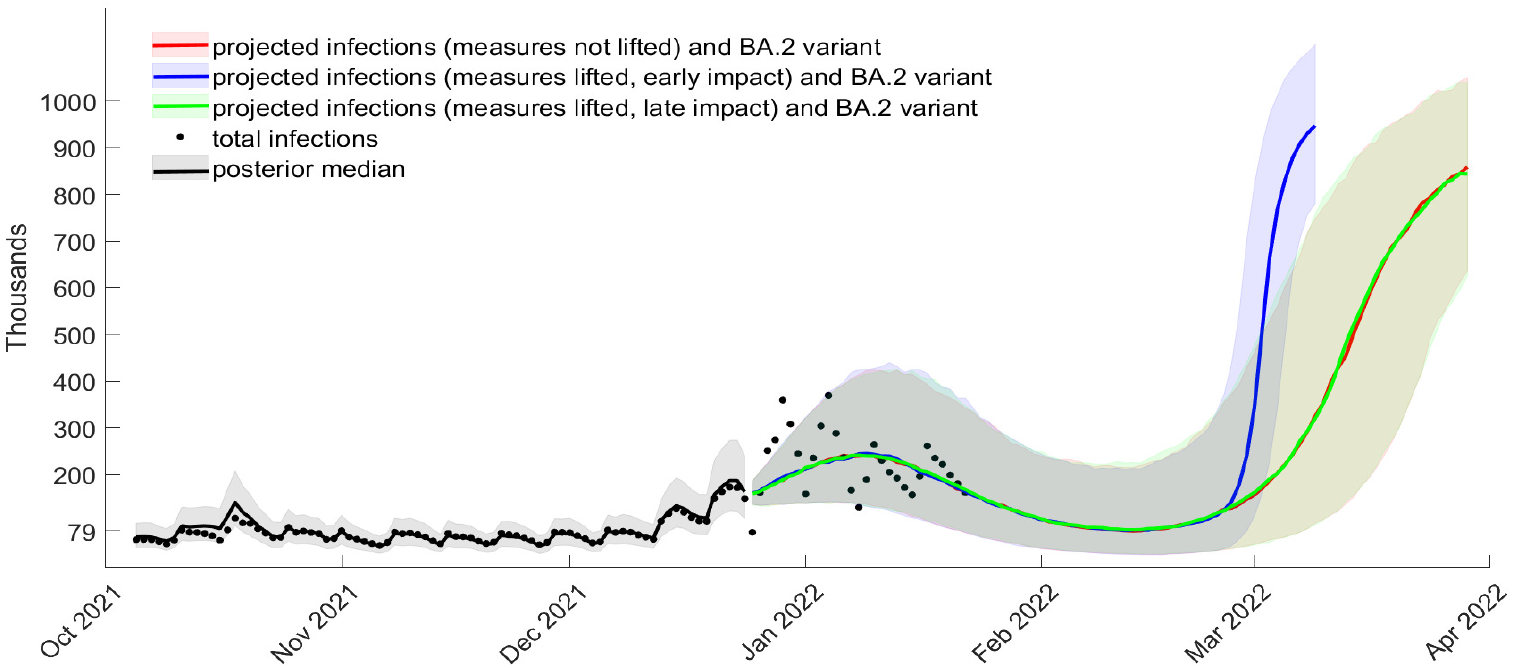
Projected total cases from December 25, 2021, waning of boosters after 5 months, *bir* = 0.75, *ρ*_*o*_ = 0.41 (posterior median), relative BA.2 intensity increase *ρ*_*BA*.2_ = 20%

### 3.5 Impact on hospitalisations

A large number of infections causes many economic disruptions, including work absences, and substantial health burdens. As a larger population is vaccinated and boosted, it may seem that the second concern becomes smaller. For this purpose, we also approximate new hospital admissions associated with the projected cases for the scenarios considered in the previous section.

Let 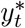 represent the median projected infections in a given scenario from Figures 10-13 and further three scenarios in the Supplementary Appendix (Section S5, Figure S21-S23) which are the same, but with the booster waning after 6 months rather than 5 months. Denote by 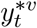 and 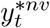 the projected infected people that had the vaccine (the second dose and the booster) and did not have the vaccine, respectively. The series for 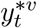 was obtained by multiplying 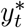 by the fraction vaccinated individuals in the total reported cases 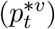. We obtained this fraction by using data from the COVID-19 vaccine weekly surveillance reports published week 48 of 2021 until week 4 of 2022. 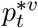 was obtained by dividing the sum of reported individuals which received a second dose more than 14 days before the specimen date plus those which received boosters by total cases minus those cases for which the booster status was unknown. Weekly values for 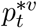 for weeks 47 to 52 of 2021 and 1 to 3 of 2022 were obtained by linear interpolation (these varied from as low as 52.71% in week 47 to 70.21% in week 51 of 2021). After January 22, 2022 we maintained 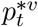 constant at 61.38%. The series for 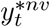 series was obtained by multiplying 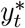 by 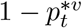.

To these series we apply a risk of hospitalisation for the Omicron BA.1 variant of 1.5551%. This was obtained as follows: *r*_*o*_ = 0.33 × 1 × 4.7% = 1.551%, where 0.33 is the hazard ratio of Omicron relative to Delta (UKHSA, 2021), 1 is the hazard ratio of Delta relative to Alpha (Veneti et al., 2022) and 4.7% is the adjusted absolute risk of hospital admission for the Alpha variant (Nyberg et al., 2021). Then, 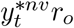 and 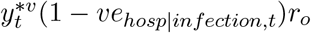 give the hospitalised infected non-vaccinated and hospitalised infected vaccinated, where *ve*_*hosp*|*infection,t*_ is the vaccine effectiveness against hospitalisation conditioned on infection which (following Viana et al., 2021, equation (14)) can be obtained from:

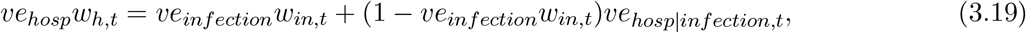

with the difference that the vaccine protection wanes, where *ve*_*infection*_ is the vaccine efficacy against infection for the Omicron BA.1 variant, *ve*_*hosp*_ is the vaccine efficacy against hospitalisation and *w*_*in,t*_ is the waning of the vaccines protection against infections. For the Omicron BA.1 variant we consider *ve*_*infection*_ = 0.7 (UKHSA, 2022, p.4) and waning of vaccine protection against infections *w*_*in,t*_ is given by (2.11) but with *t*^+^ = February 14, 2022 (when the vaccine booster protection in the projections in Figures 10-12 starts waning). We consider *ve*_*hosp*_ = 0.95 (UKHSA, 2022, p.8) which wanes over time. The waning of the vaccine protection against hospitalisations is assumed *w*_*h,t*_ = *w*_*in,t*_. It follows from (3.19)

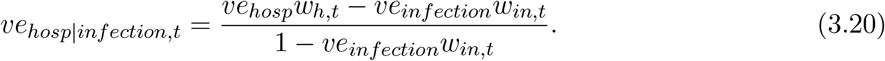

Our projections for new hospital admissions start on January 2, 2022 (projected infections start on 25 of December, 2021 but we assume a delay in hospital admissions for the Omicron BA.1 variant of 7 days, see ONS, 2021). Figure 14 shows the projected new admission into hospital based on the median of projected infections from Figures 10-13 when waning of the vaccine booster is after 5 months (in Section S6 of the Supplementary Appendix we present the projected new admissions into hospital assuming the waning of boosters is after 6 months). The figure also includes observed new hospital admissions (by date of hospitalisation) for England until January 22, 2022 (obtained from the official Coronavirus in the UK dashboard).

**Figure 14:**
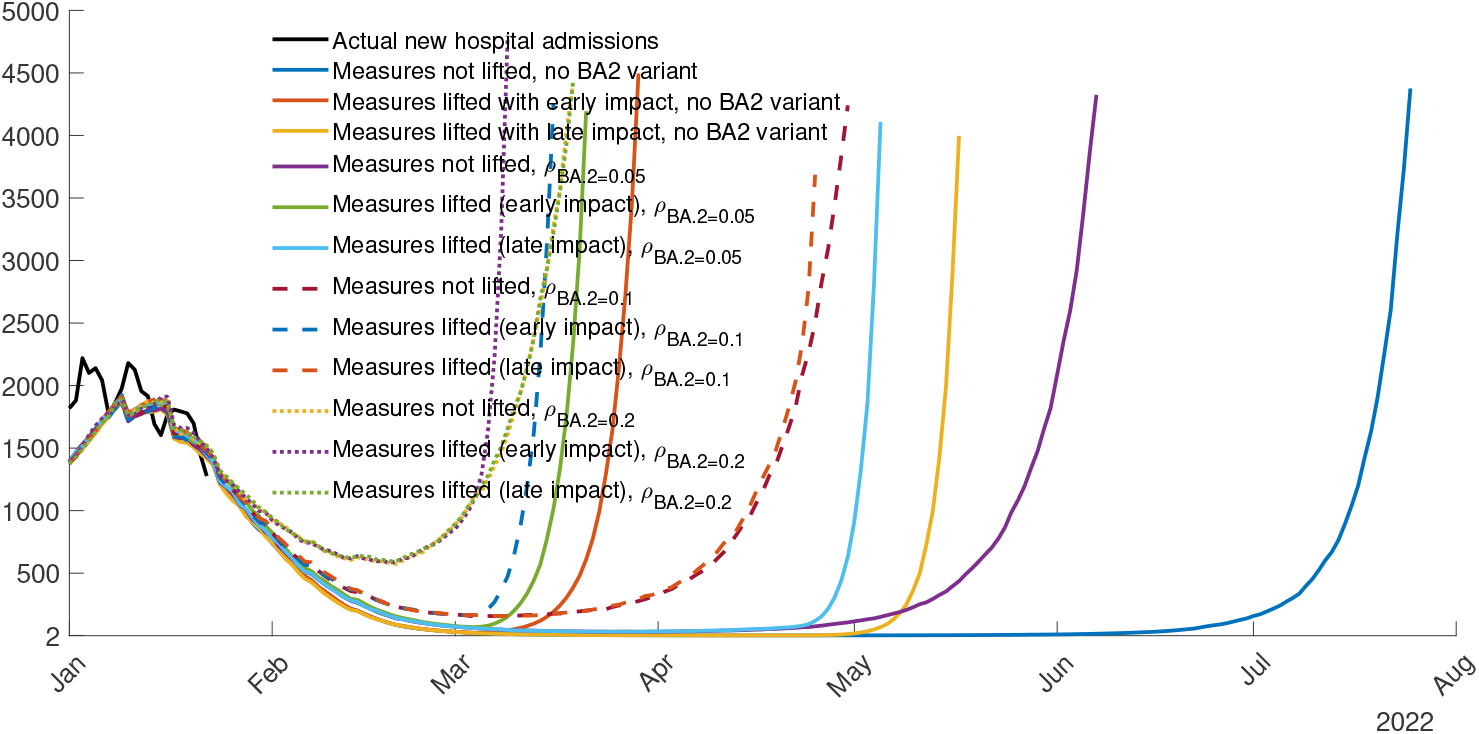
Projected new admissions into hospital based on the median projected infections from Figures S13-S15, waning of boosters after 5 months, *bir* = 0.75, *ρ*_*o*_ = 0.41 (posterior median)

As can be seen from Figure 14, the projected new hospital admissions match well actual new hospital admissions in the period for which observations are available. The projections show that with a new infection wave, new hospitalisations rise steeply and quickly reach the peak of new hospital admissions experienced during the pandemic so far (4,134 new hospital admissions on January 12, 2021). It is possible that these projections are too large because we did not model temporary immunity protection conferred by a recent infection, which would require a more complex epidemiological setup. In that case, Figure 14 is still relevant for policy makers in approximating the timing of a rise of hospital admissions.

## 4 Conclusion

We proposed a dynamic intensity model for SARS-CoV-2 infections in England to disentangle between NPIs, vaccines uptake and variants of concern.

We find that NPIs were effective at reducing infections in all waves so far, but that they worked best with the wild-type variant, which is natural given the fact that more infectious variants are harder to contain. We also found that the decrease in effectiveness of the same NPIs due to more infectious variants was strongly mitigated by vaccines and boosters.

Our counterfactuals show that had the booster campaign started one month earlier or if it had reached faster a significant fraction of the population then the winter wave in December 2021 could have been avoided. We also show that a two week lockdown implemented early would have been much more effective at reducing infections in December 2021 than the longer semi-lockdown actually implemented.

Projections for the next few months of 2022 from the estimate model show that, as booster protection wanes, another wave is predicted to occur. The predicted timing for the new wave is affected by several factors: 1) NPIs; 2) infectiousness of Omicron BA.2 variant; 3) timing for the waning of booster protection; and 4) effectivity of boosters at reducing infection intensity. Our analysis also reveals that, whenever a new wave of infections is projected to occur in a given scenario, new hospital admissions increase substantially shortly afterwards.

Even though our analysis is tailored to England, the framework we developed can be used for any country for which total cases can be inferred, and data on variants and vaccines is available. While our scenarios are focused on Omicron, our framework can also be employed for new variants of concern, to inform policy makers about the necessity and timing of further booster campaigns and non-pharmaceutical interventions.

## Data Availability

All data is publicly available and the source of data is mentioned in the manuscript.

## Supplementary Appendix

### S1. Ratio of reported to total cases

Figure S1 shows the time-variation in total to reported cases, interpolated linearly every two weeks, constructed assuming reported cases have a two-day or a five-day delay from infectiousness to reporting.

**Figure S1:**
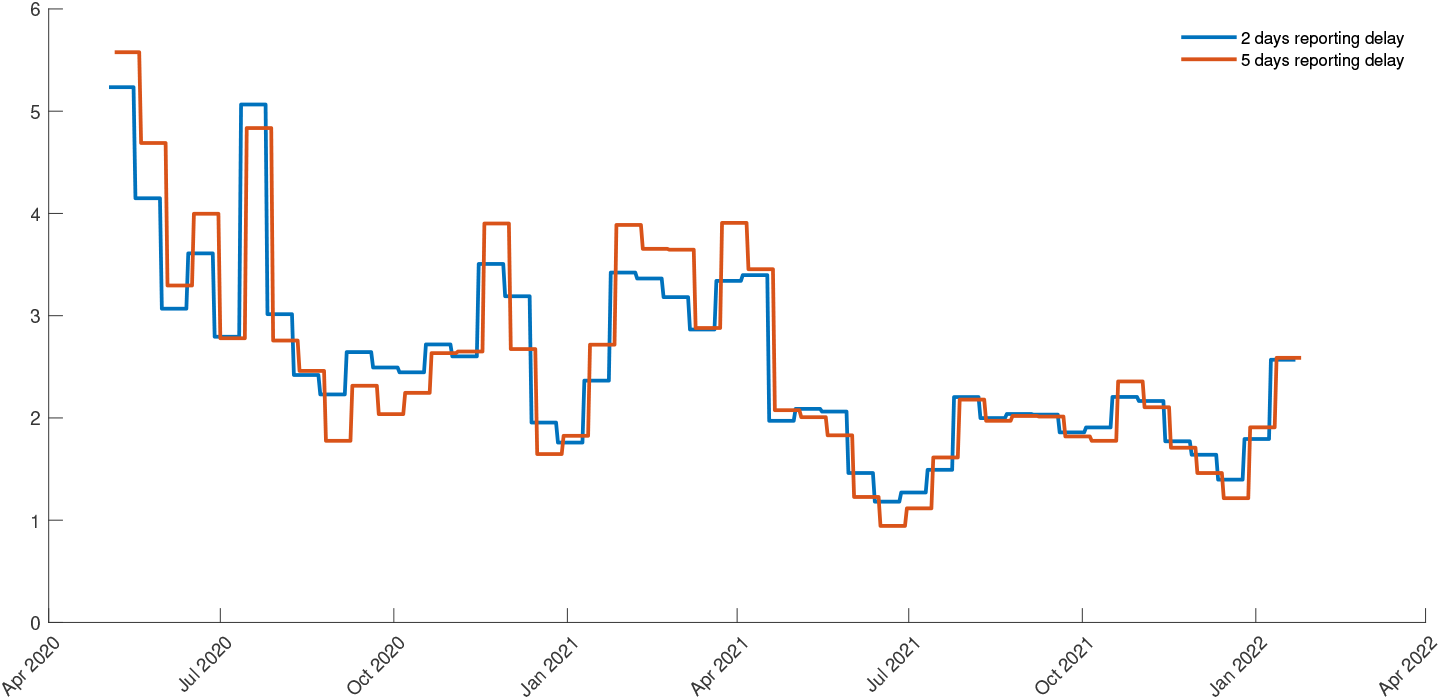
Ratio of total cases to reported cases

### S2. Posteriors and Identification

Figures S2-S8 plot the posterior distributions (and the posterior median) of all parameters against their priors. Here, recall that the parameters, *θ*_*i*_ ≥ 0 and *β*_*i*_ ≥ 0, *i* = 0, …, 9, associated with *y*_*t*−1_ and 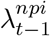 change in each regime by *γ*_*i*_ and *ω*_*i*_ respectively: *θ*_*i*_ = *θ*_*i*−1_ + (−1)^*i*^*γ*_*i*_, *β*_*i*_ = *β*_*i*−1_ + (−1)^*i*^*ω*_*i*_ *i* = 1, …, 9, *γ*_*i*_ = *θ*_*i*_ − *θ*_0_, and *ω*_*i*_ = *β*_*i*_ − *β*_0_, *i* = 1, …, 9.

We note that, while some parameters enter in our model specification as products (see (2.3) in the main paper), they are in fact identified over different periods, often non-overlapping or only partially overlapping. For example, 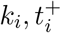 (the steepness and midpoint parameters in the NPIs transition functions) are identified in their own regime *i* where other transition functions are already fixed (the transition functions for the variants-of-concern, the vaccine 2nd dose and booster *g*_*i,t*_, *i* = *α, δ, o, v, b*). Moreover, *θ*_0_, *β*_0_, *γ*_*i*_, *ω*_*i*_, *i* = 1, …, 9 (associated with the previous day infections *y*_*t*−1_ and previous daily intensity *λ*_*t*−1_) are also identified in their own regime which do not fully overlap with samples over which *ρ*_*α*_, *ρ*_*δ*_, *ρ*_*o*_, *vir, bir* are identified. However, there are parameters that are identified only by very volatile periods or short periods, such as the parameters corresponding to the second and to the sixth regime of the NPIs (describing the transition from some relaxation measures to the second lockdown in November 2021, and the transition to relaxations and the Euro 2020 football tournament).

Therefore, to further check identification, we fixed the parameters at their posterior median, generated 100 samples from the model, re-estimated the model on each of these samples, and displayed in Figures S2-S8 the fraction of medians that are outside the 90% credible range (this Bayesian identification analysis is recommended in Aitchison, 1962); see the values of *z* in Figures S2-S8. As expected, for most parameters, this fraction is below 10%, as should be the case if the parameters are identified, and there are only two regime parameters that seem less well identified: *ω*_2_ and *ω*_6_ which correspond *λ*_*t*−1_ in the second and the sixth regime. Additionally, we note that the posterior density of the booster intensity reduction parameter *bir* has a large overlap with the prior, perhaps because the sample over which this parameter is identified is too short.

**Figure S2:**
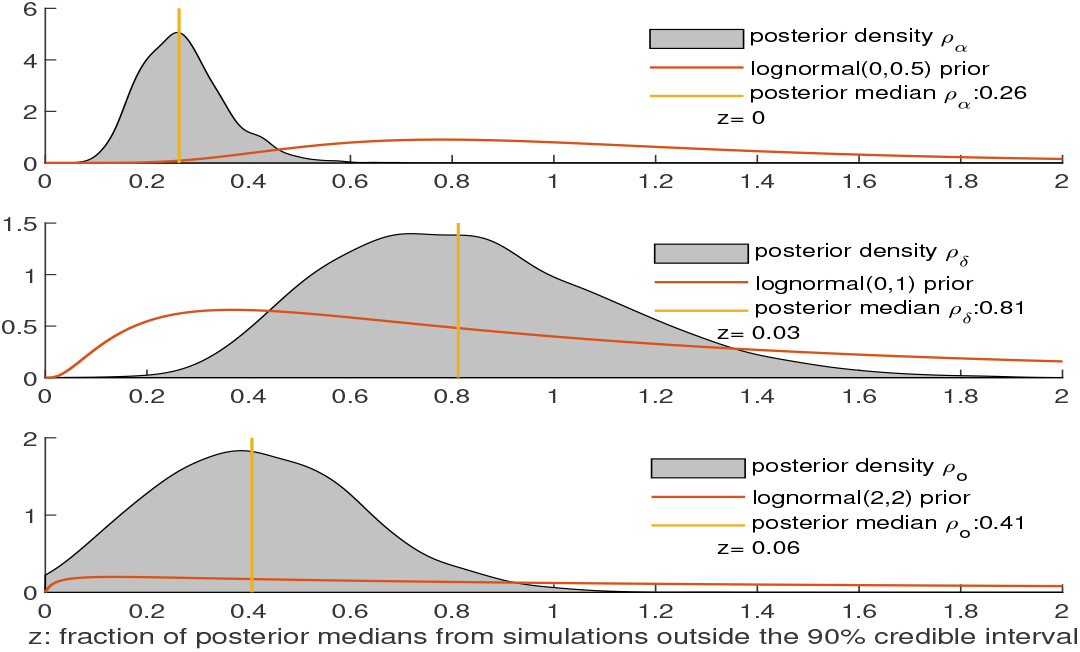
Prior and posterior for *ρ*_*α*_, *ρ*_*δ*_ and *ρ*_*o*_

**Figure S3:**
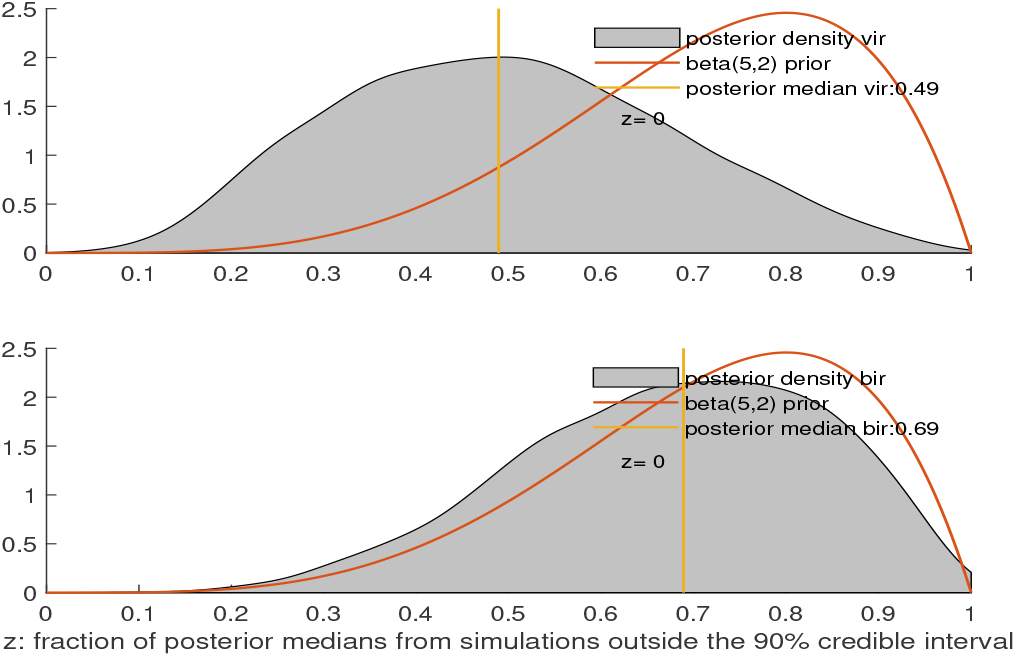
Prior and posterior for *vir* and *bir*

**Figure S4:**
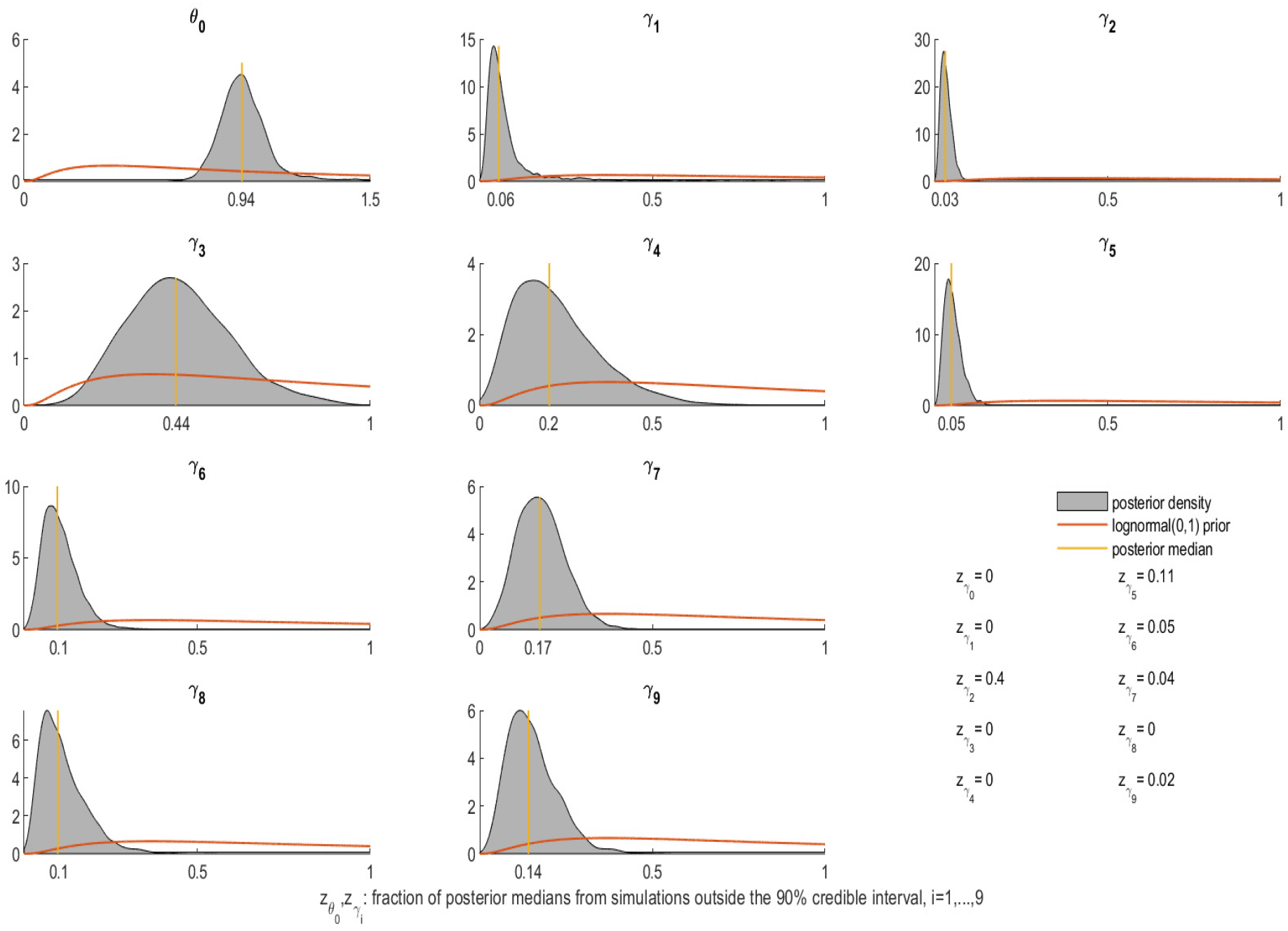
Prior and posterior for *θ*_0_ and *γ*_*i*_ *i* = 1, …, 9

**Figure S5:**
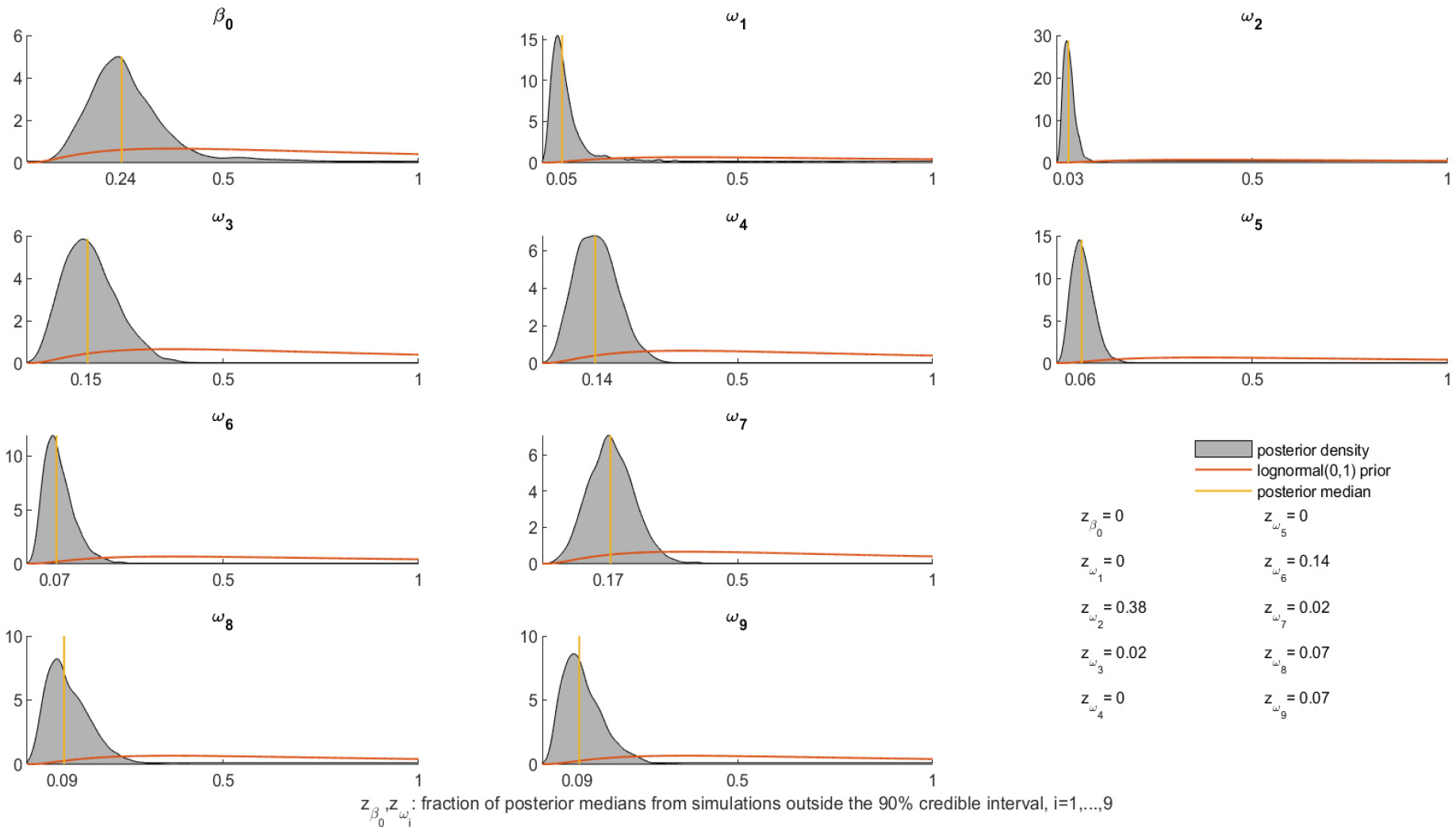
Prior and posterior for *β*_0_ and *ω*_*i*_ *i* = 1, …, 9

**Figure S6:**
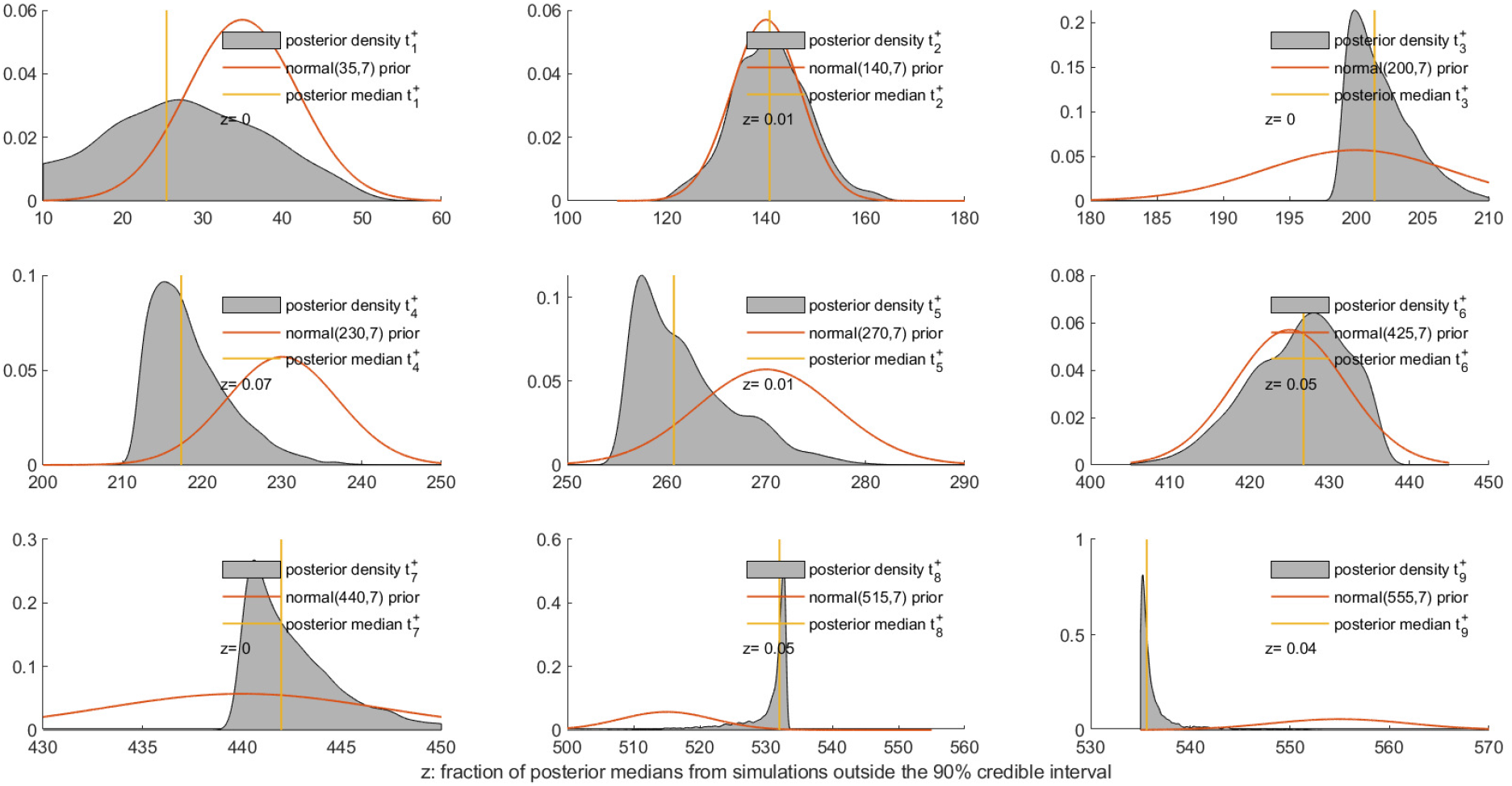
Prior and posterior for 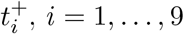

**Figure S7:**
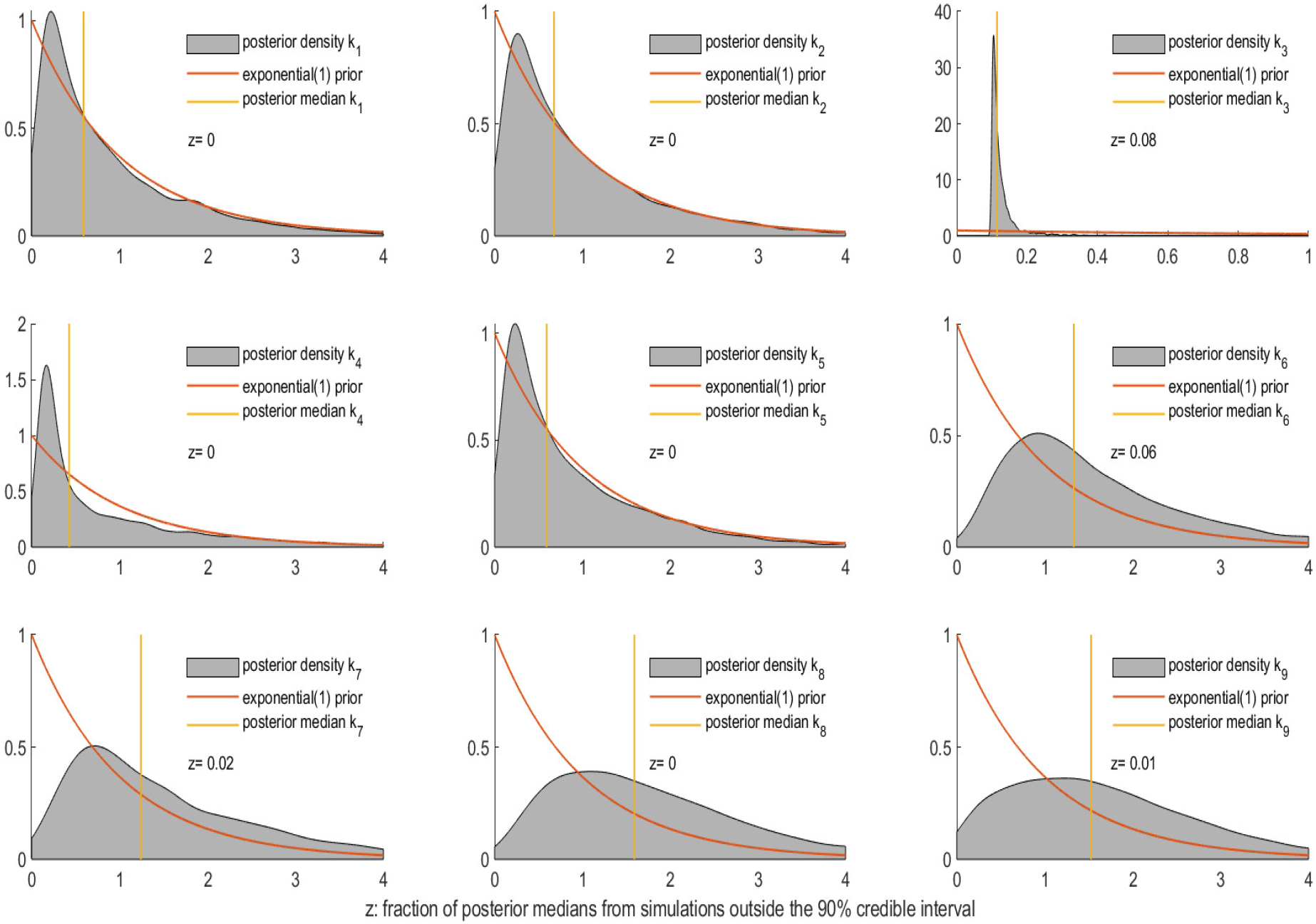
Prior and posterior for *k*_*i*_, *i* = 1, …, 9

**Figure S8:**
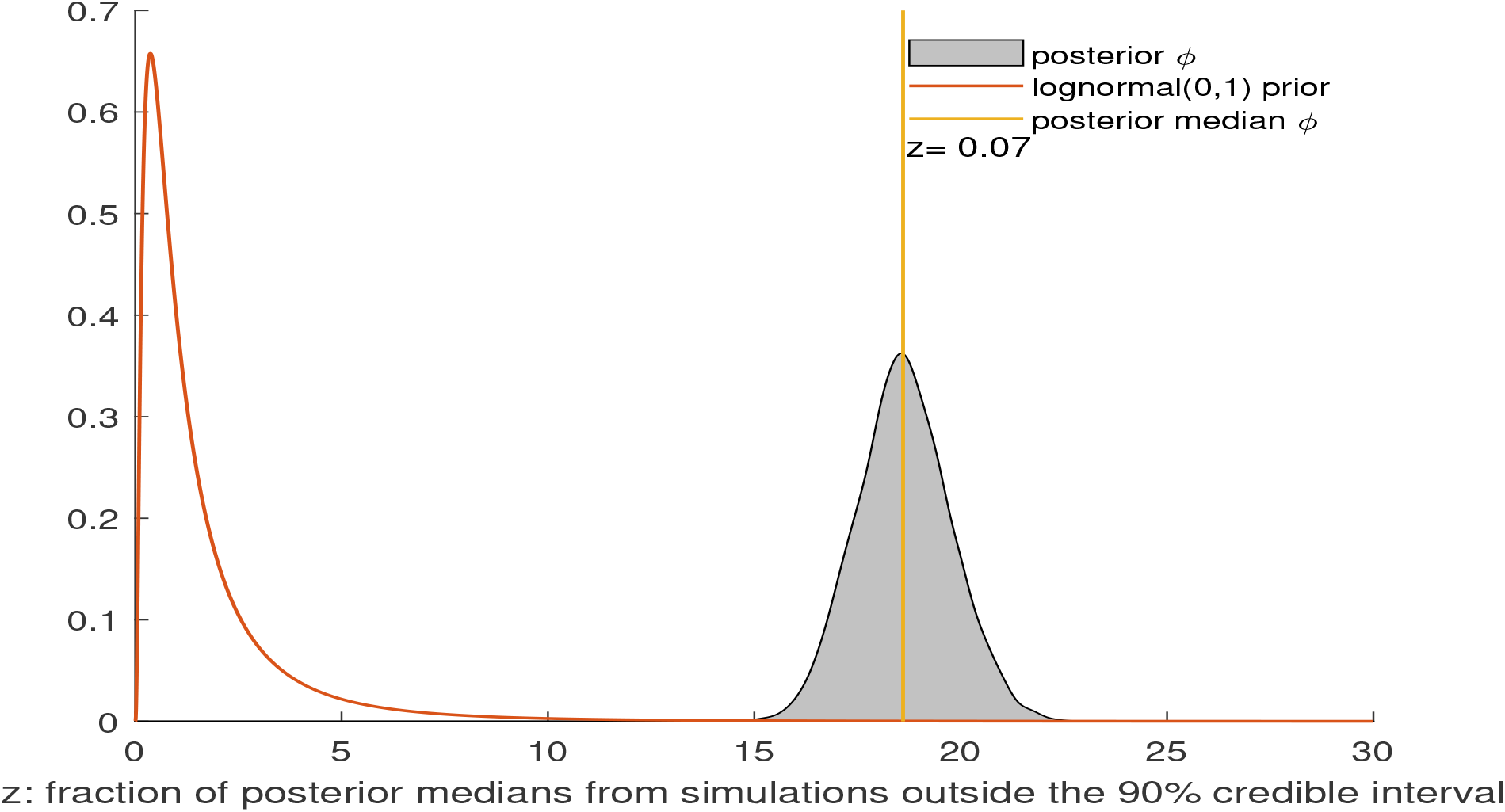
Prior and posterior for *ϕ*

### S3. Counterfactuals in Section 3.3 repeated with the lower 5% to the upper 95% quantiles

**Figure S9:**
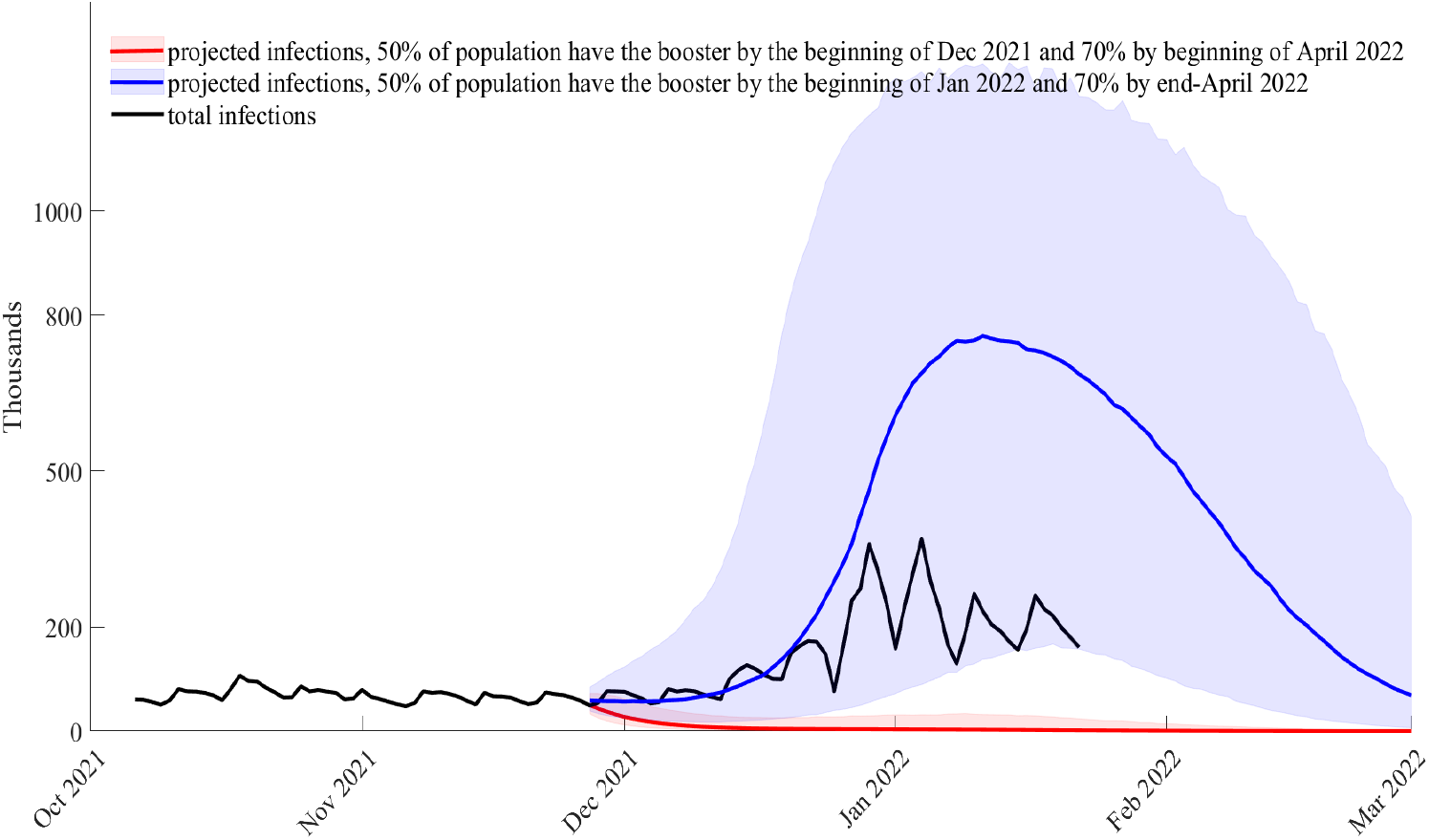
Counterfactual when the vaccine booster campaign starts on August 16, 2021, and the population is reached faster (red) or slower (blue); projection of daily infections from November 27, 2021; *bir* = 0.69 (posterior median) and *ρ*_*o*_ = 0.41 (posterior median)

**Figure S10:**
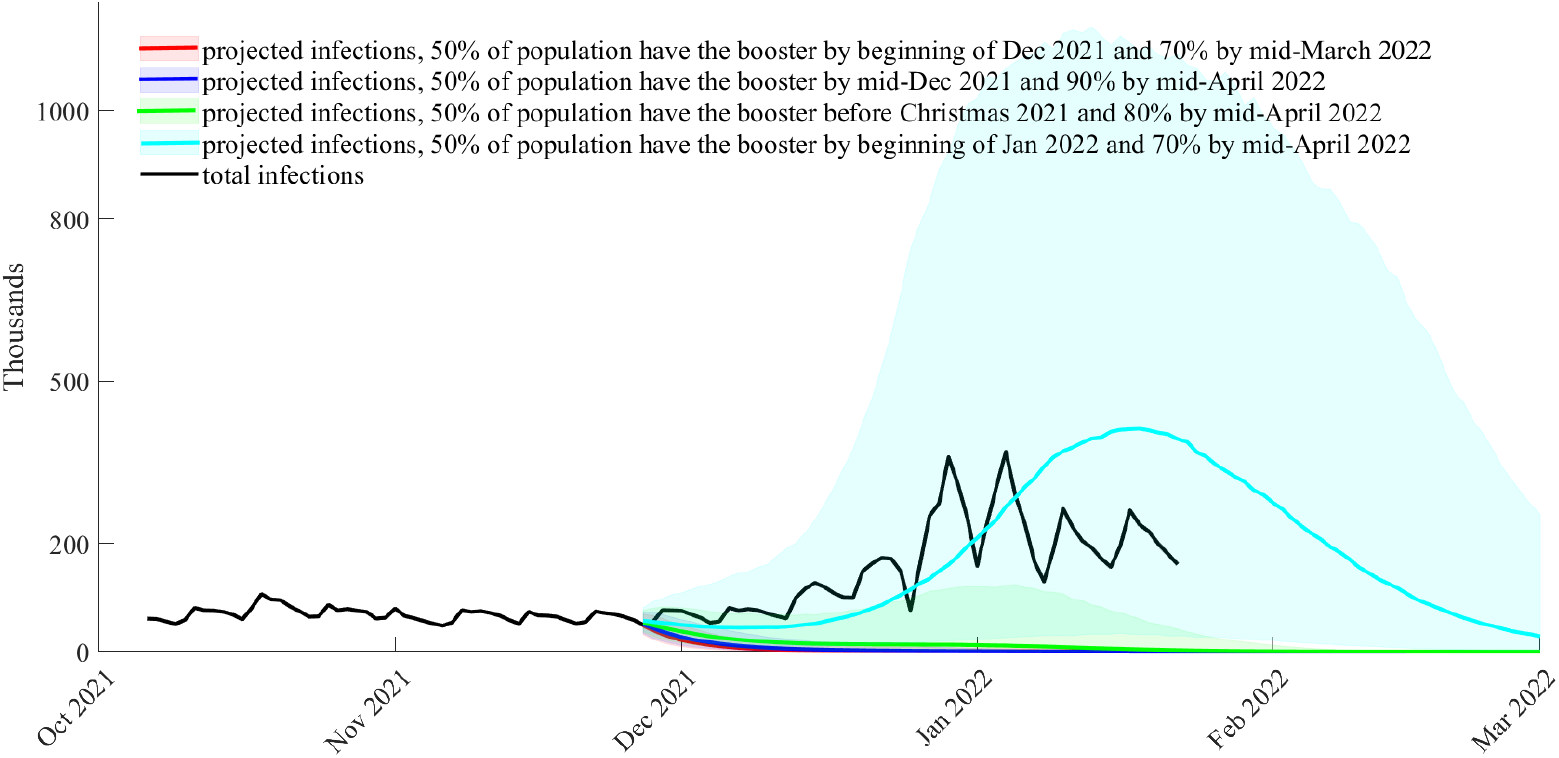
Counterfactual when the vaccine booster campaign starts on September 16, 2021, and population is reached at different speeds; projection of daily infections from November 27, 2021; *bir* = 0.69 (posterior median) and *ρ*_*o*_ = 0.41 (posterior median)

**Figure S11:**
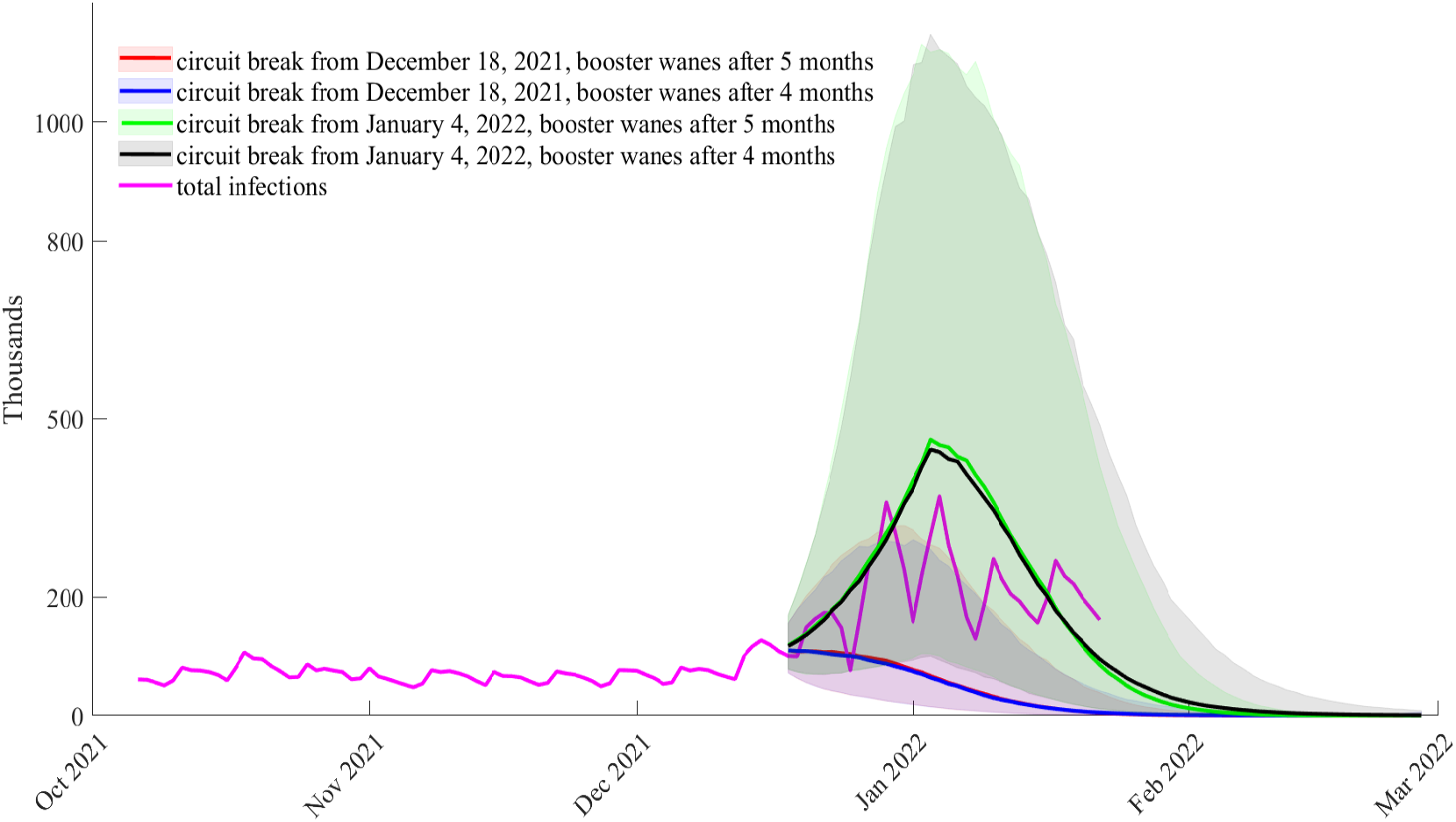
Counterfactual when there is a circuit break (2 weeks hard lockdown) from December 18, 2021, or January 4, 2022 (peak of infections); projection of daily infections from December 18, 2021; *bir* = 0.69 (posterior median) and *ρ*_*o*_ = 0.41 (posterior median)

**Figure S12:**
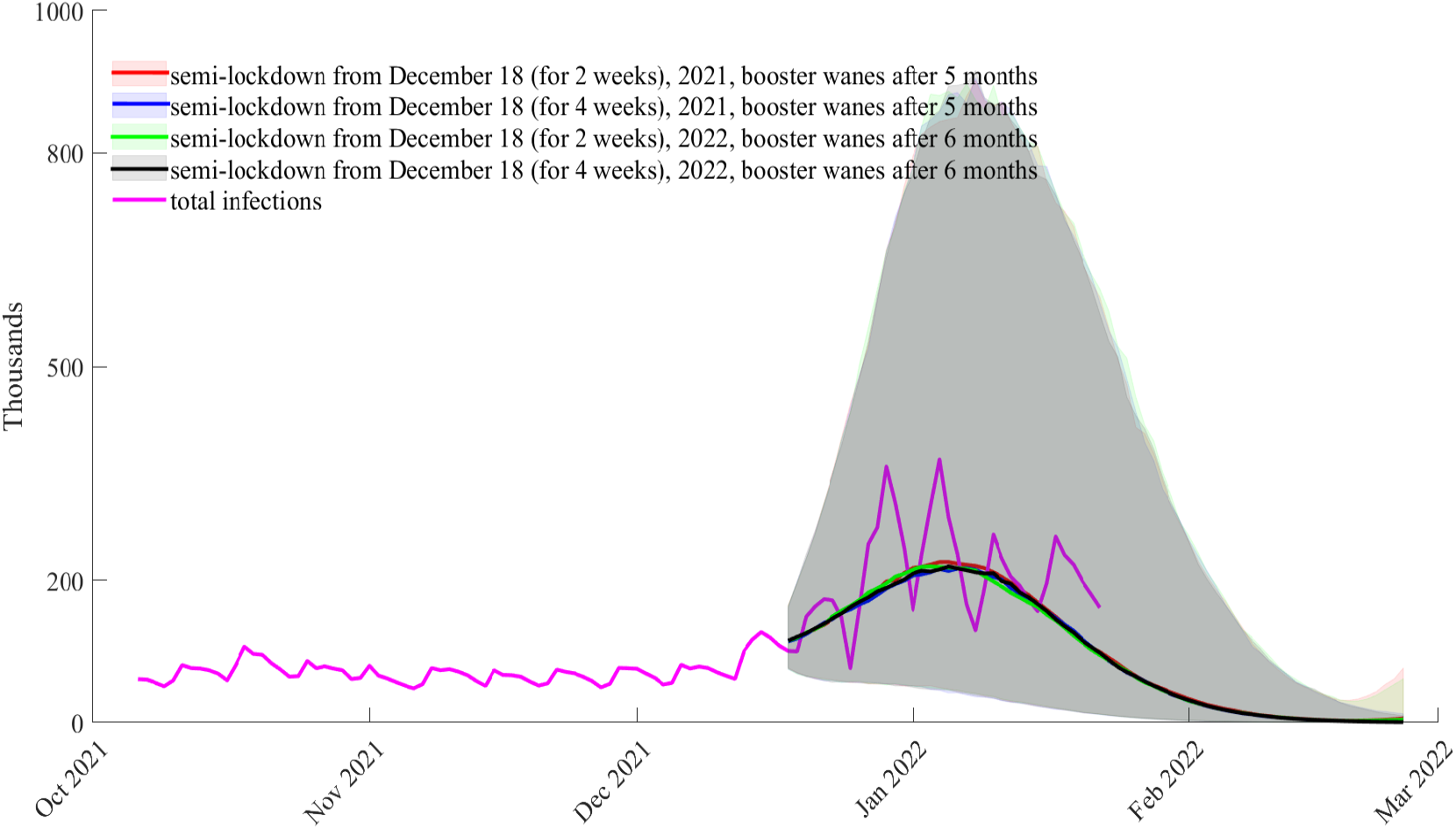
Counterfactual when there is a semi-lockdown from December 18, 2021; projection of daily infections from December 18, 2021; *bir* = 0.69 (posterior median) and *ρ*_*o*_ = 0.41 (posterior median)

### S4. Scenarios in Section 3.4 repeated with the lower 5% to the upper 95% quantiles

**Figure S13:**
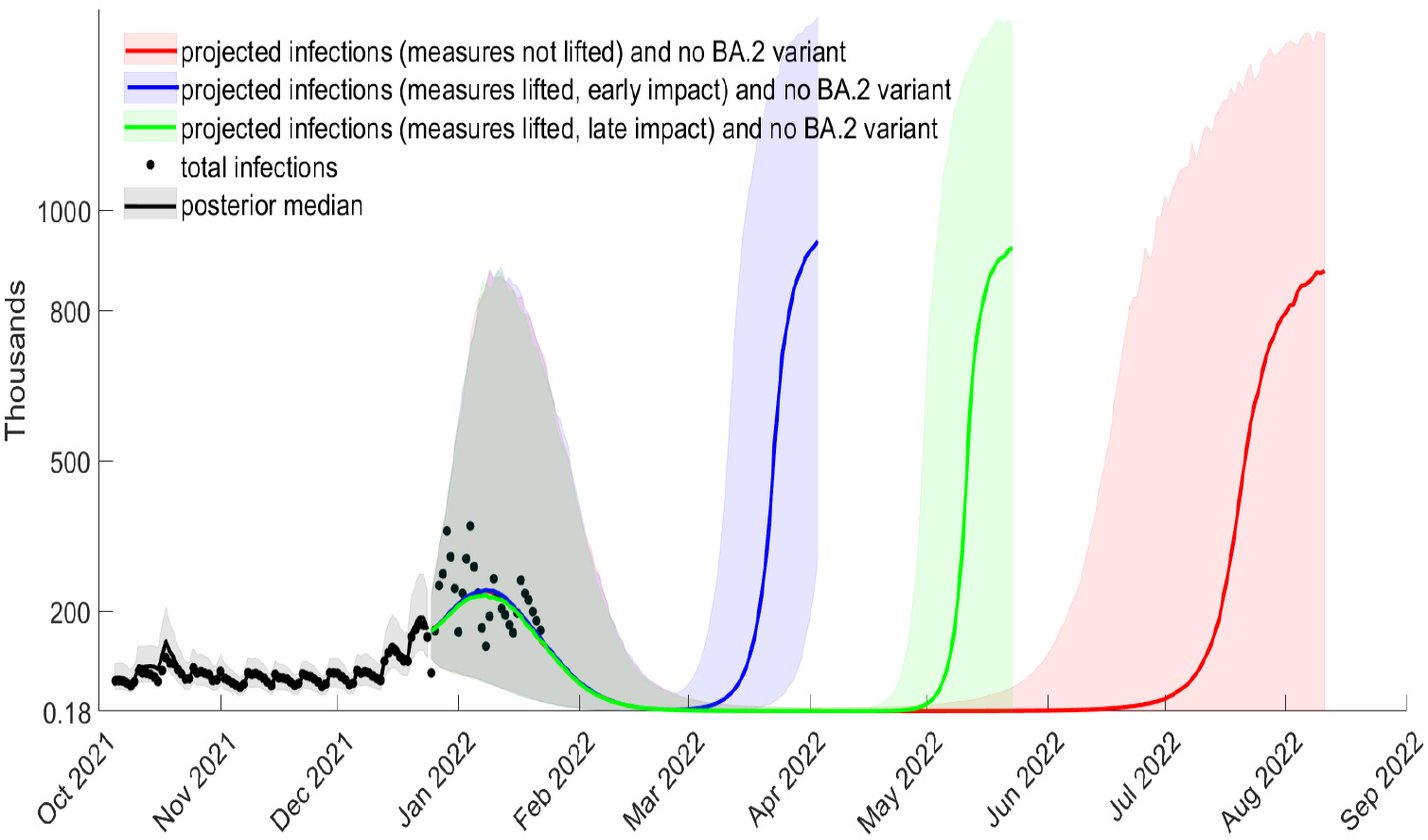
Projected total cases from December 25, 2021, waning of boosters after 5 months, *bir* = 0.75, *ρ*_*o*_ = 0.41 (posterior median), no increase in relative BA.2 intensity

**Figure S14:**
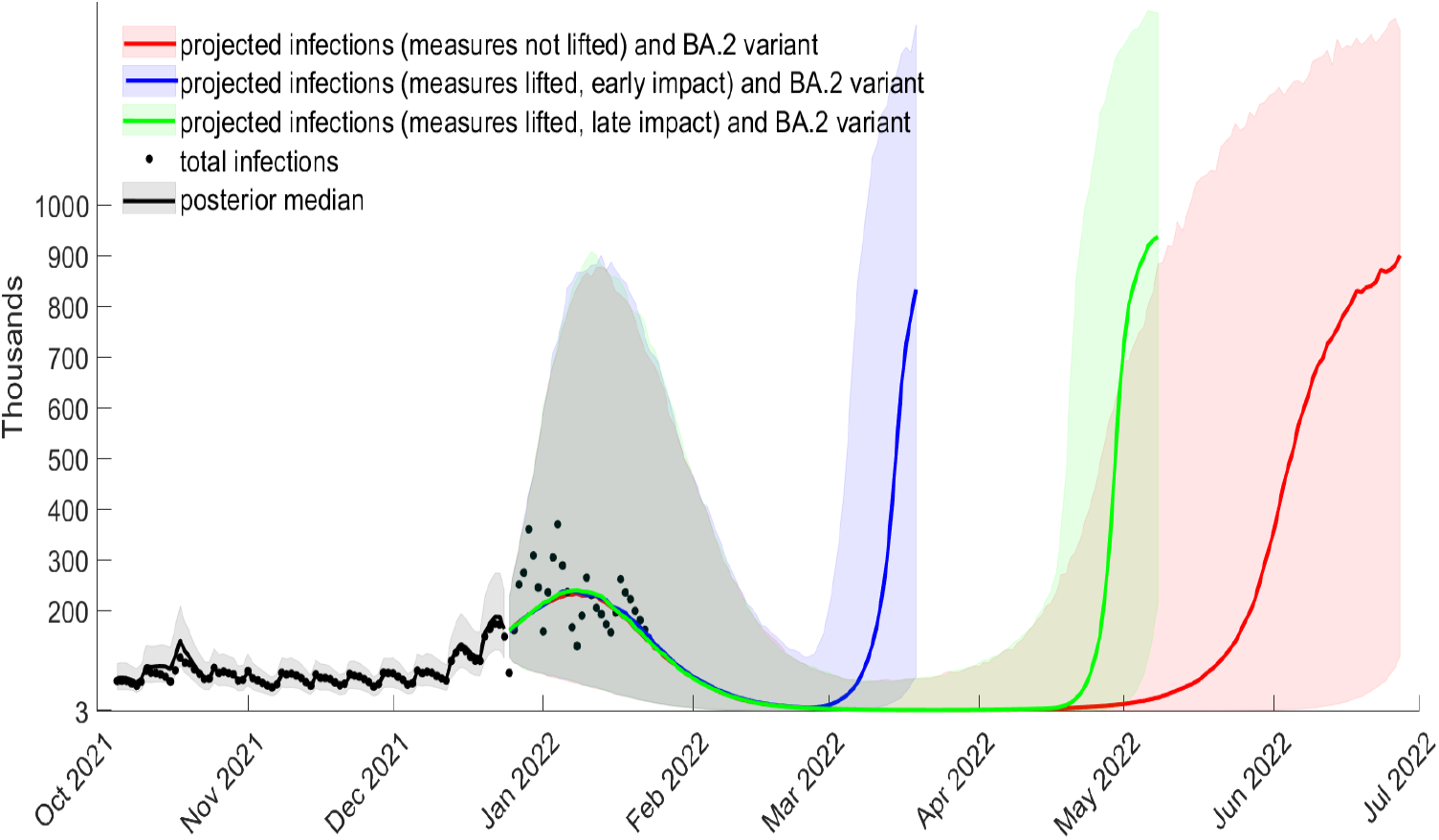
Projected total cases from December 25, waning of boosters after 5 months, *bir* = 0.75, *ρ*_*o*_ = 0.41 (posterior median), relative BA.2 intensity increase *ρ*_*BA*.2_ = 5%

**Figure S15:**
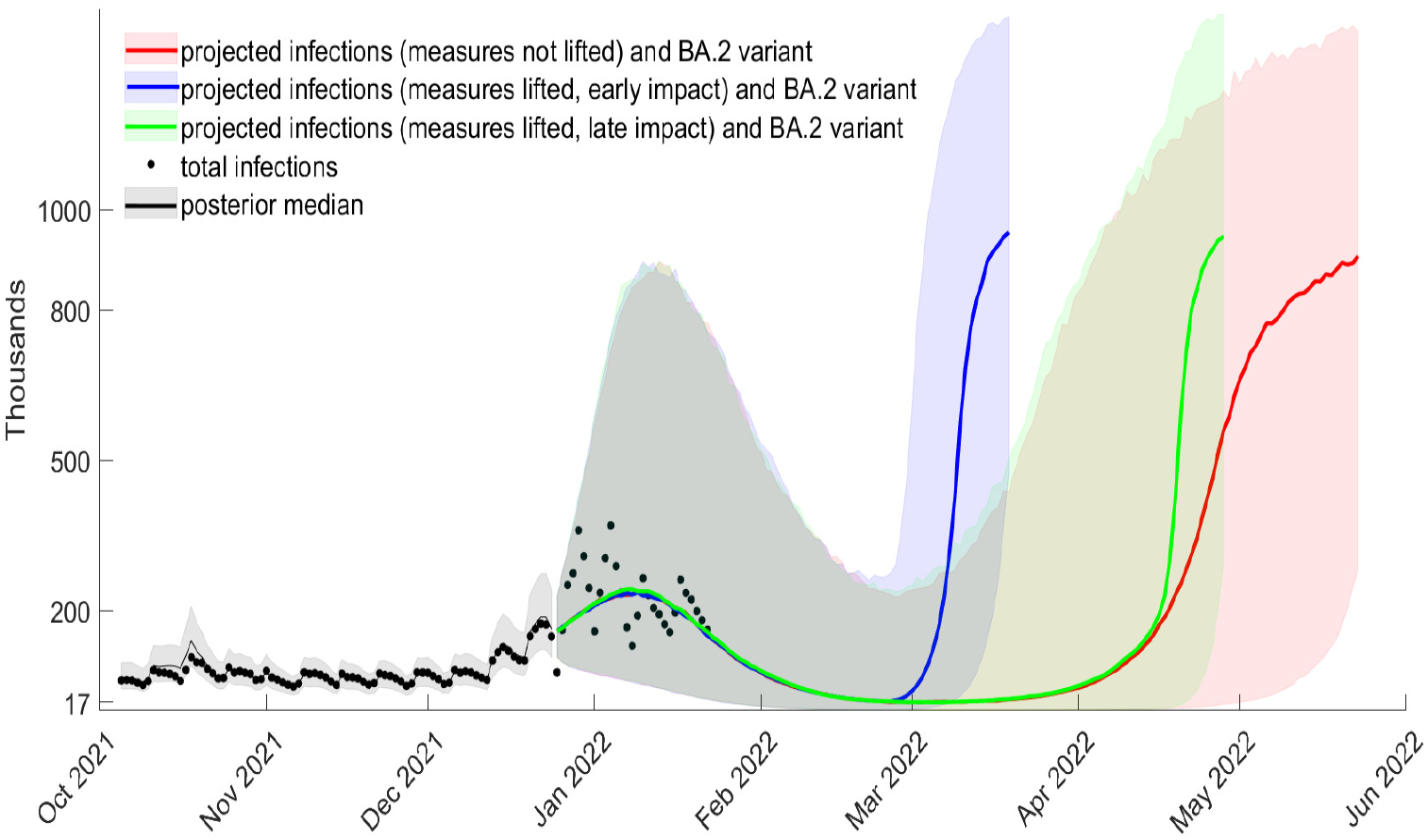
Projected total cases from December 25, 2021, waning of boosters after 5 months, *bir* = 0.75, *ρ*_*o*_ = 0.41 (posterior median), relative BA.2 intensity increase *ρ*_*BA*.2_ = 10%

**Figure S16:**
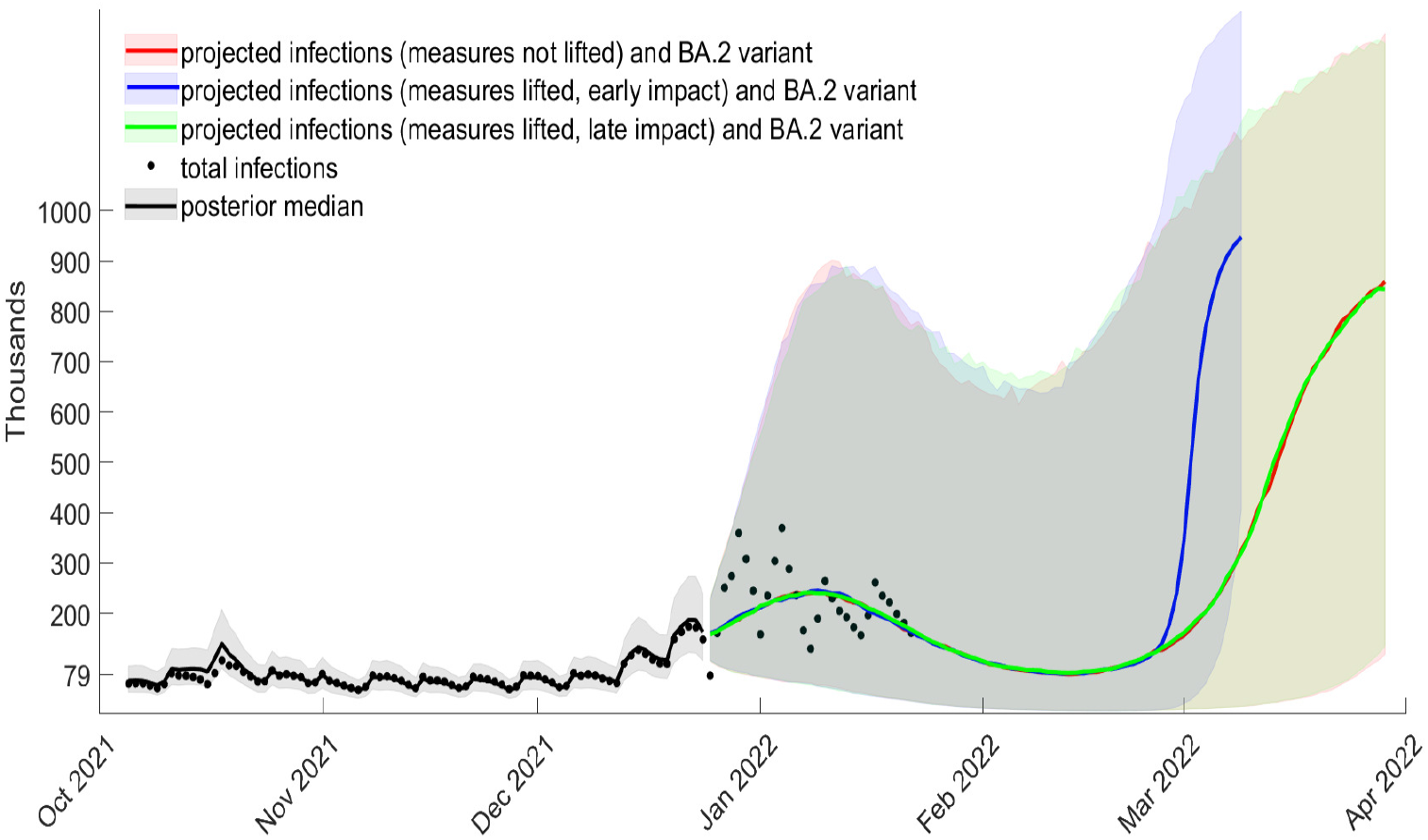
Projected total cases from December 25, 2021, waning of boosters after 5 months, *bir* = 0.75, *ρ*_*o*_ = 0.41 (posterior median), relative BA.2 intensity increase *ρ*_*BA*.2_ = 20%

### S5. Further scenarios for boosters and their waning

In Figures S17 and S18 (with the interquartile range) we look at what happens if the booster intensity reduction is only 0.69 rather than the 0.75 assumed so far in the main paper (Section 3.4). In this case a projected new wave happens earlier (compared to the case when *bir*=0.75). The figures are repeated in Figures S19 and S20 with the lower 5% to the upper 95% quantiles.

**Figure S17:**
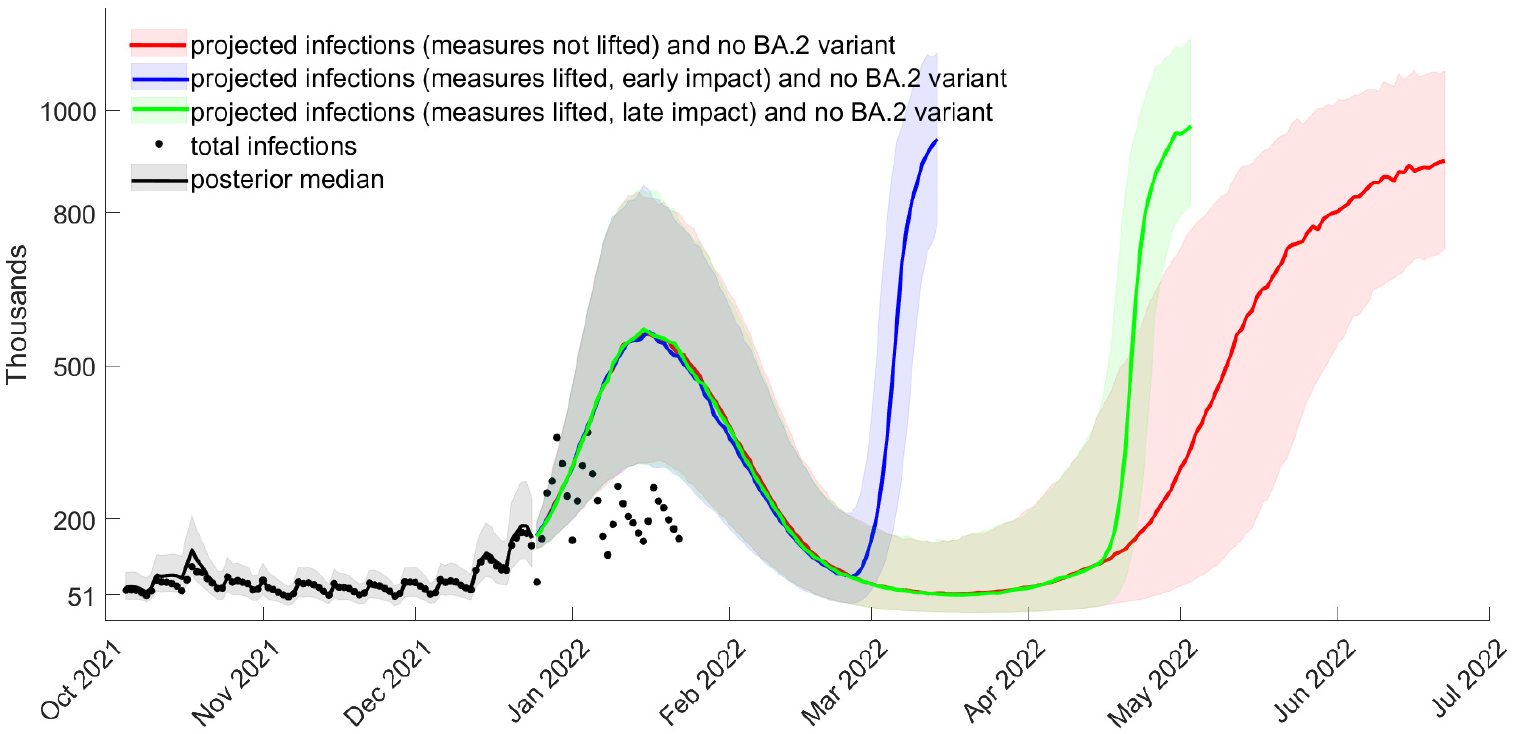
Projected total cases from December 25, 2021, waning of boosters after 5 months, *bir* = 0.69 (posterior median), *ρ*_*o*_ = 0.41 (posterior median), with no BA.2 variant, with the interquartile range

**Figure S18:**
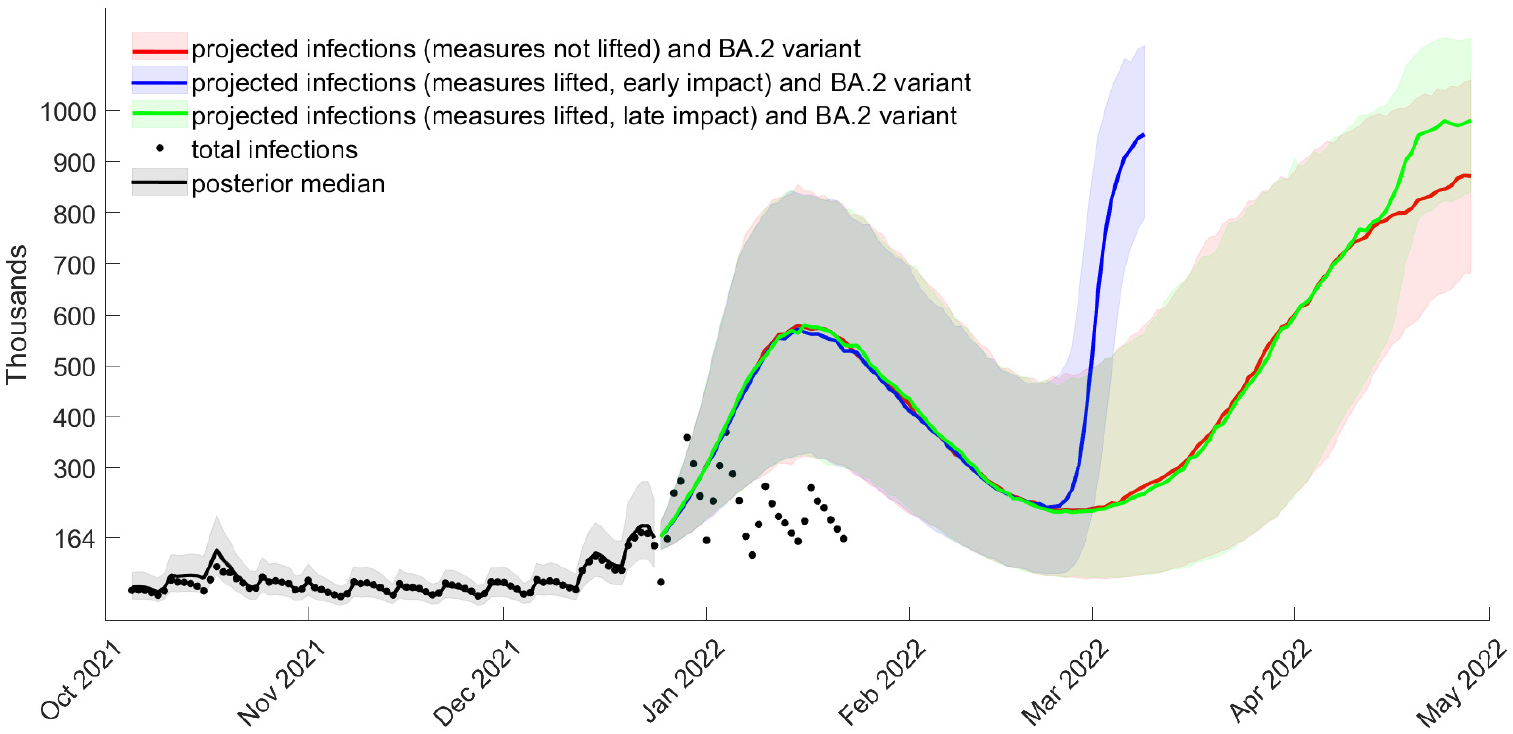
Projected total cases from December 25, 2021, waning of boosters after 5 months, *bir* = 0.69 (posterior median), *ρ*_*o*_ = 0.41 (posterior median), relative BA.2 intensity increase *ρ*_*BA*.2_ = 0.05, with the interquatile range

**Figure S19:**
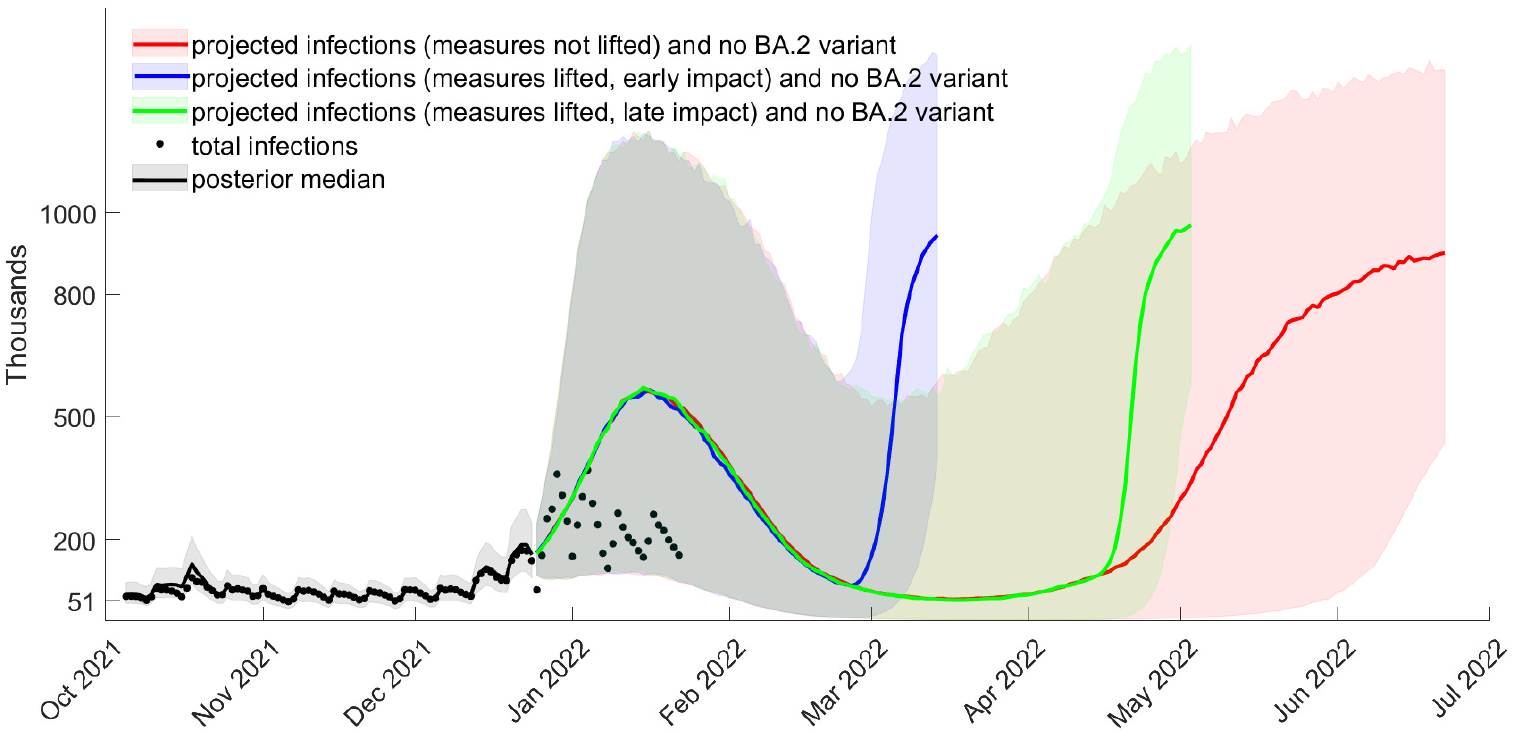
Projected total cases from December 25, 2021, waning of boosters after 5 months, *bir* = 0.69 (posterior median), *ρ*_*o*_ = 0.41 (posterior median), with no BA.2 variant, with the lower 5% to the upper 95% quantiles

**Figure S20:**
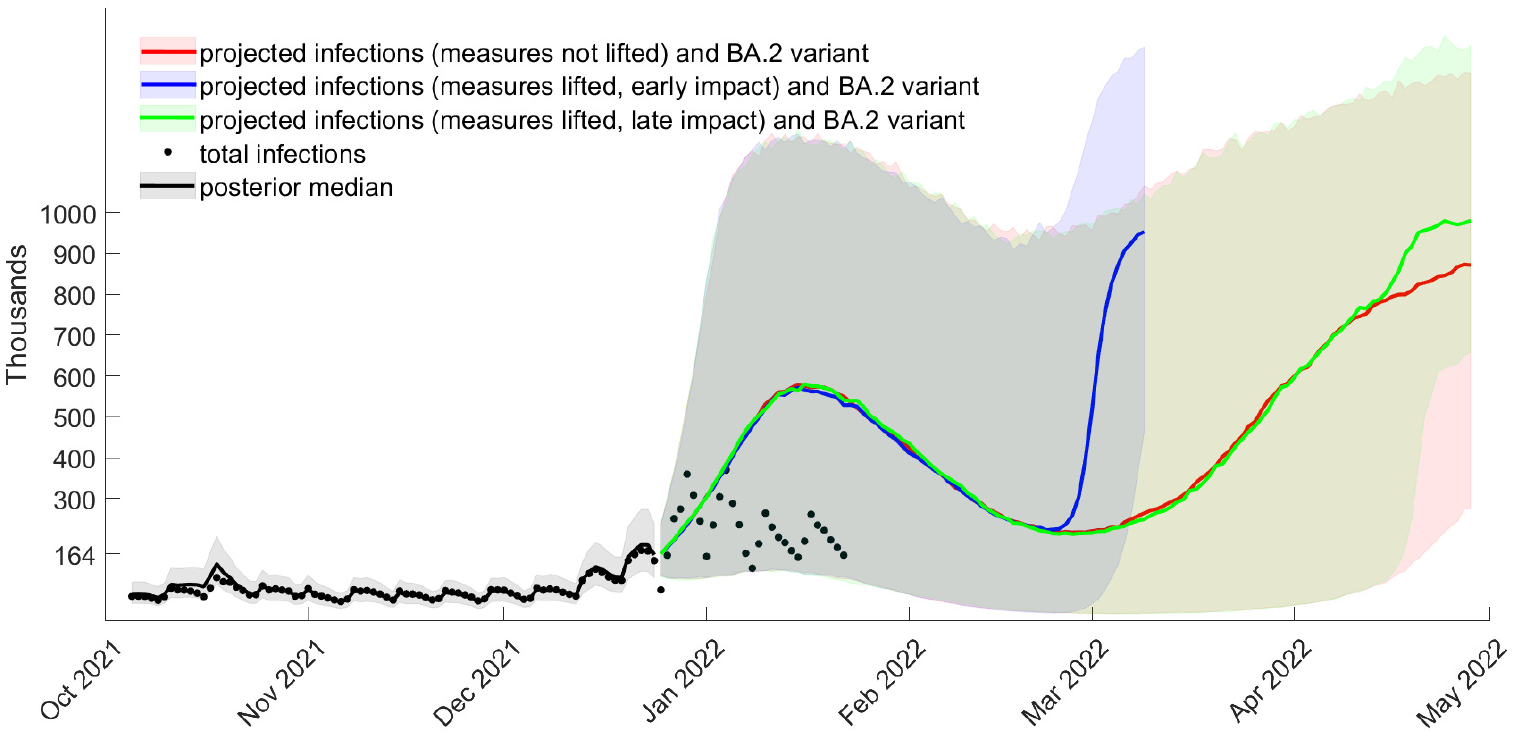
Projected total cases from December 25, 2021, waning of boosters after 5 months, *bir* = 0.69 (posterior median), *ρ*_*o*_ = 0.41 (posterior median), and relative BA.2 intensity increase *ρ*_*BA*.2_ = 0.05, with the lower 5% to the upper 95% quantiles

Figures S21-S23 show a more optimistic scenario compared to the one in the main paper (Section 3.4), in which the booster intensity reduction is still 0.75, but it wanes slower (in 6 months compared to 5 months). The figures report the interquartile range. The same figures are repeated in Figures S24-S26, but with the lower 5% to the upper 95% quantiles. We see that in most cases, an infection wave still occurs with high probability, but is substantially delayed if measures are lifted and have a late impact. Only if *ρ*_*BA*.2_ is 5%, we note that no wave occurs within the next months.

**Figure S21:**
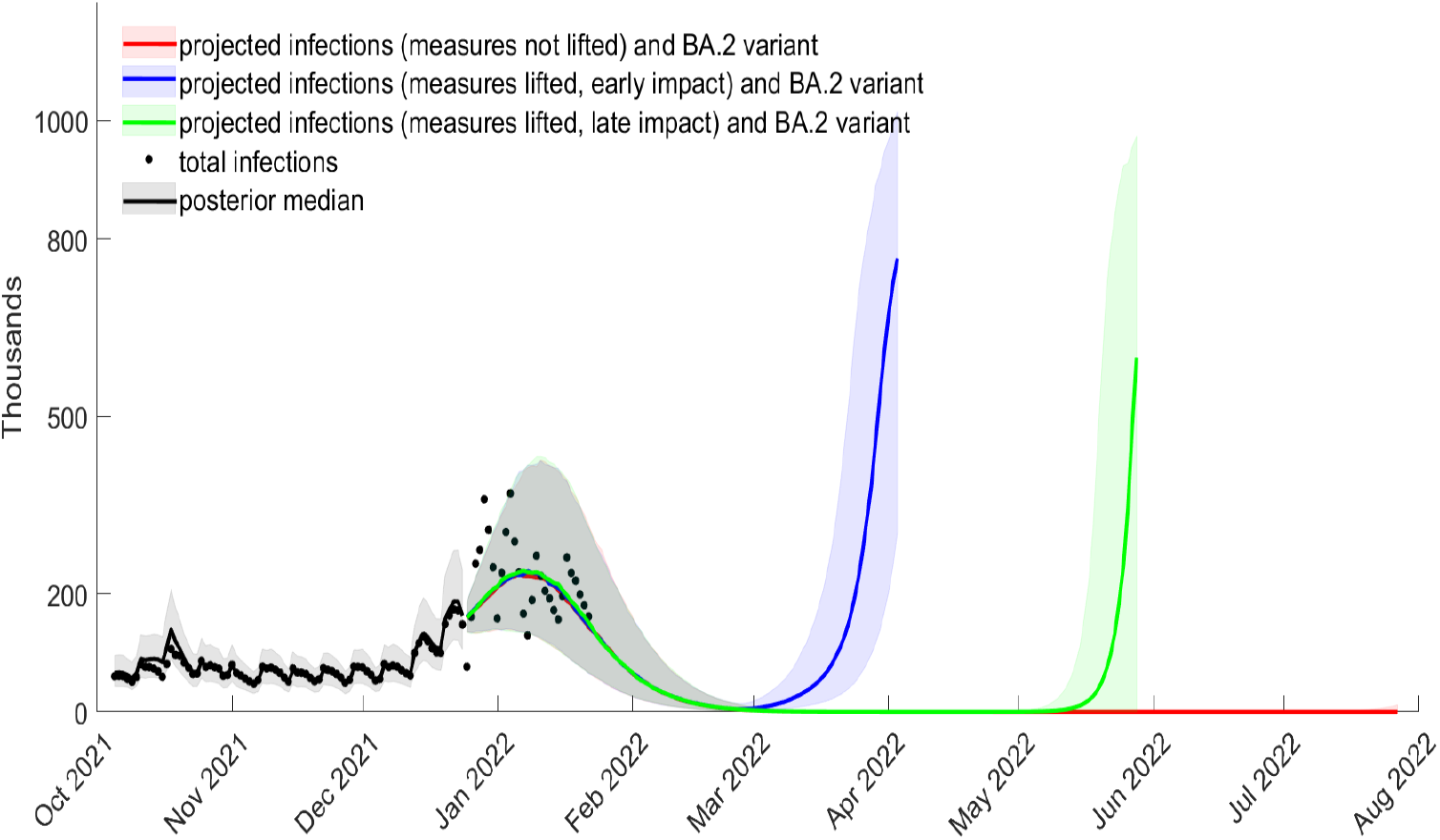
Projected total cases from December 25, 2021, waning of boosters after 6 months, *bir* = 0.75, *ρ*_*o*_ = 0.41 (posterior median), relative BA.2 intensity increase *ρ*_*BA*.2_ = 5%, with the interquartile range

**Figure S22:**
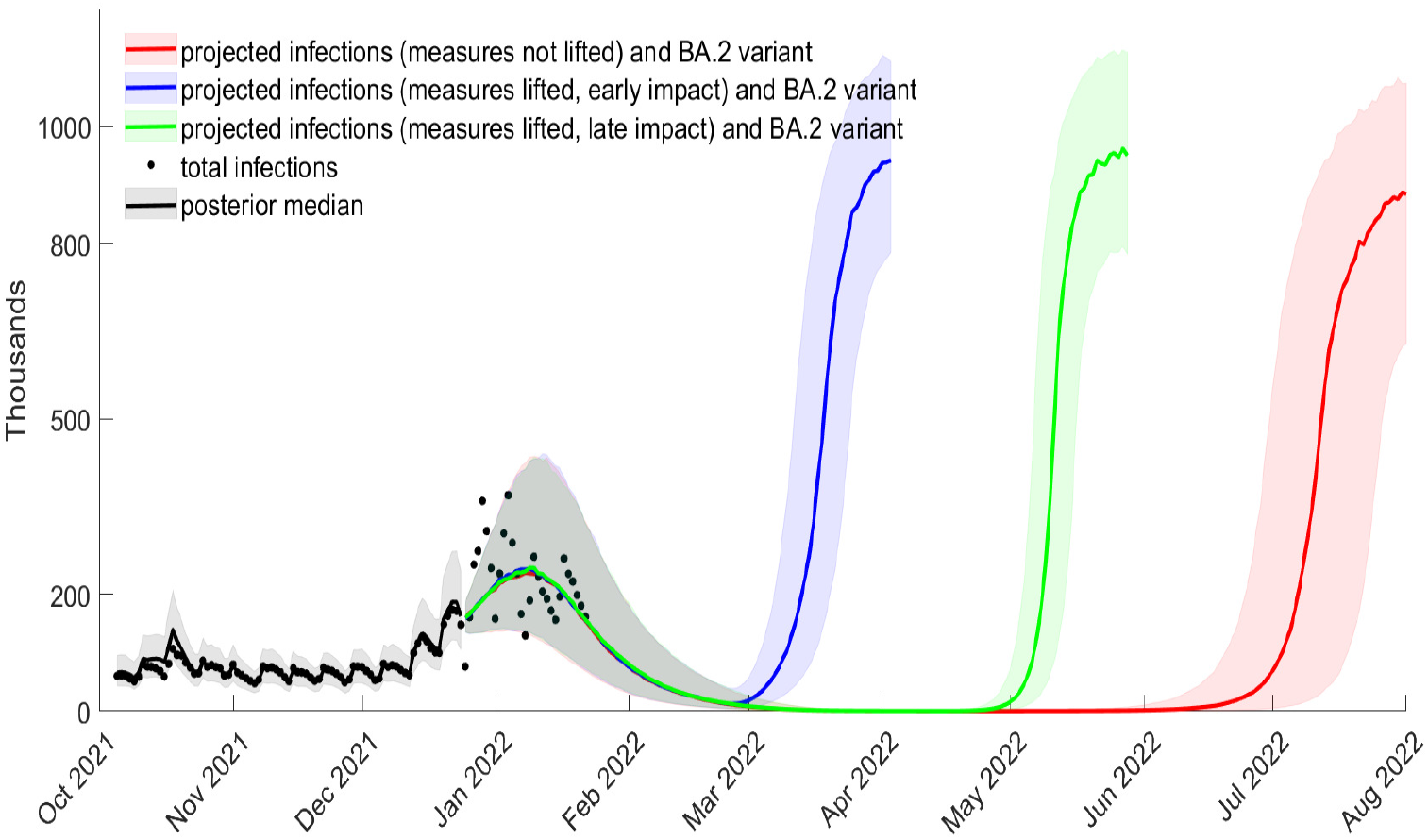
Projected total cases from December 25, 2021, waning of boosters after 6 months, *bir* = 0.75, *ρ*_*o*_ = 0.41 (posterior median), relative BA.2 intensity increase *ρ*_*BA*.2_ = 10%, with the interquartile range

**Figure S23:**
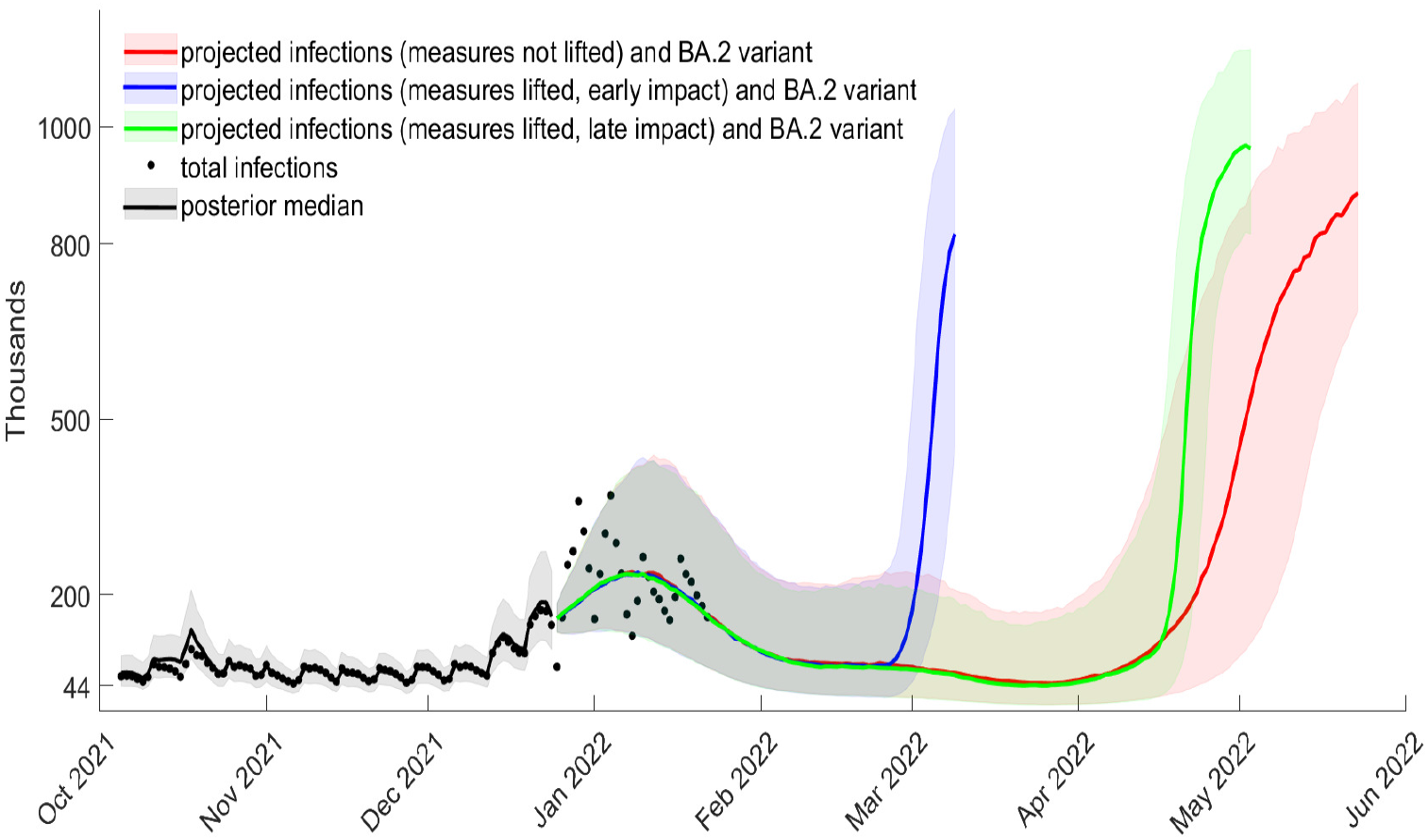
Projected total cases from December 25, 2021, waning of boosters after 6 months, *bir* = 0.75, *ρ*_*o*_ = 0.41 (posterior median), relative BA.2 intensity increase *ρ*_*BA*.2_ = 20%, with the interquartile range

**Figure S24:**
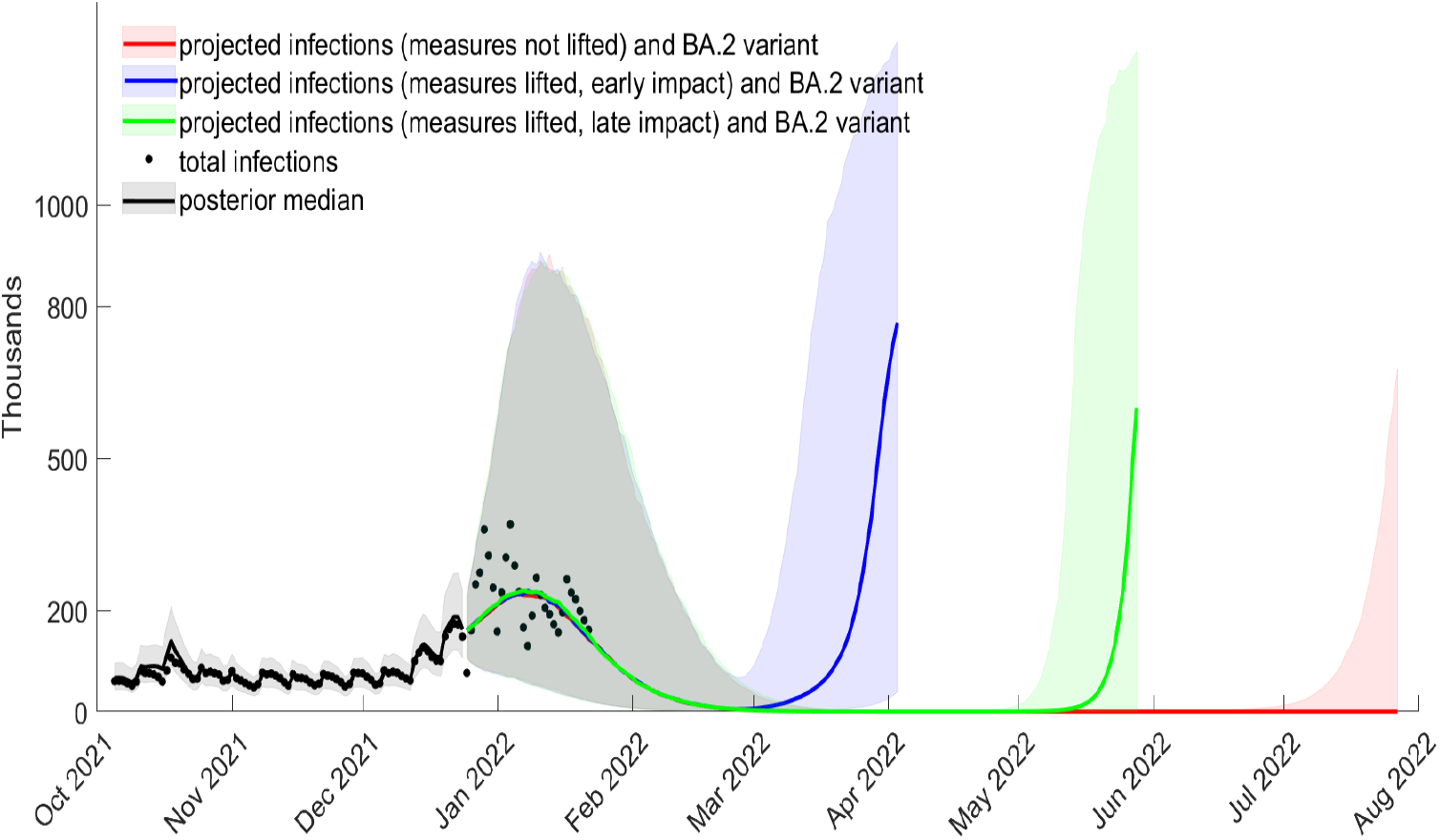
Projected total cases from December 25, 2021, waning of boosters after 6 months, *bir* = 0.75, *ρ*_*o*_ = 0.41 (posterior median), relative BA.2 intensity increase *ρ*_*BA*.2_ = 5%, with the lower 5% to the upper 95% quantiles

**Figure S25:**
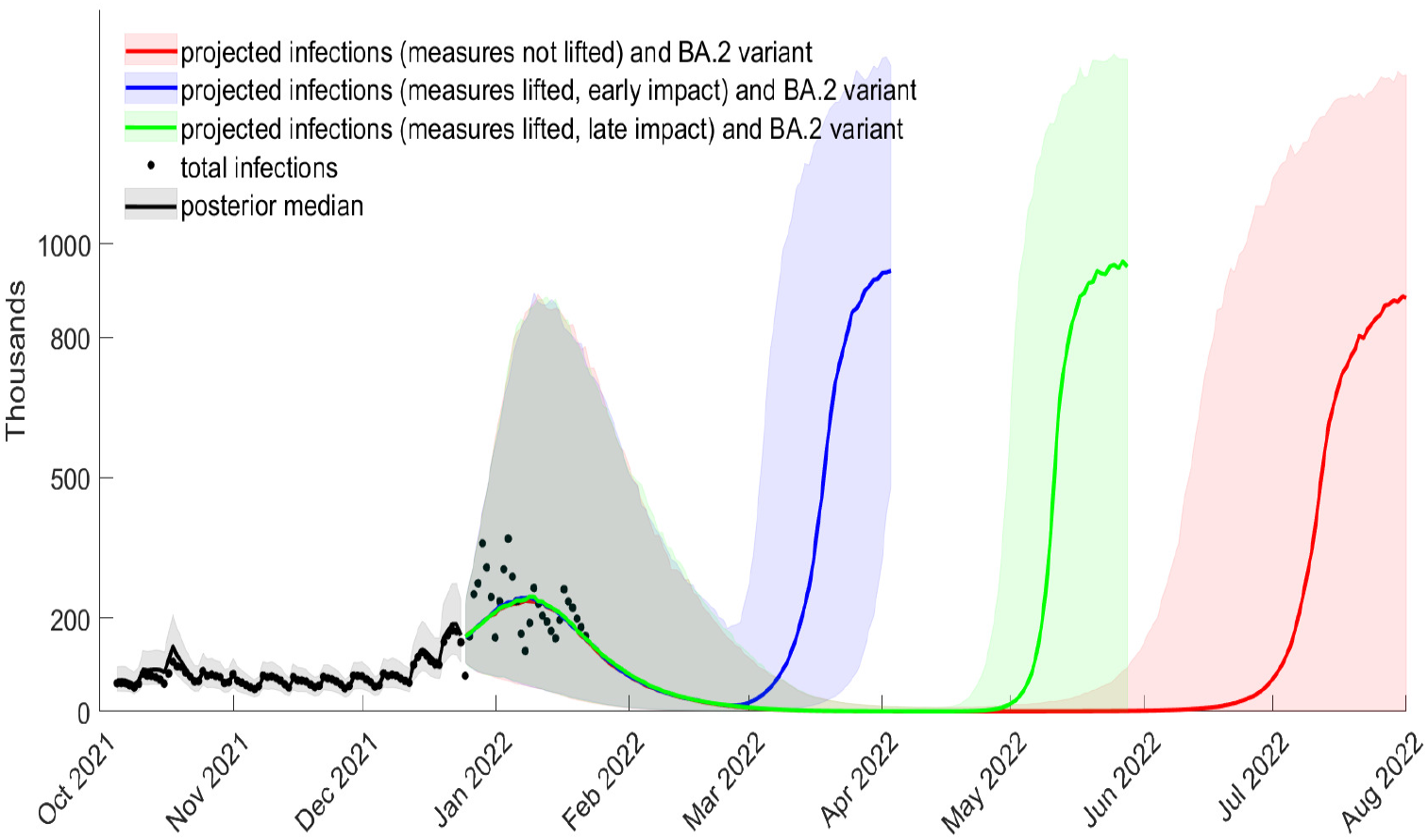
Projected total cases from December 25, 2021, waning of boosters after 6 months, *bir* = 0.75, *ρ*_*o*_ = 0.41 (posterior median), relative BA.2 intensity increase *ρ*_*BA*.2_ = 10%, with the lower 5% to the upper 95% quantiles

**Figure S26:**
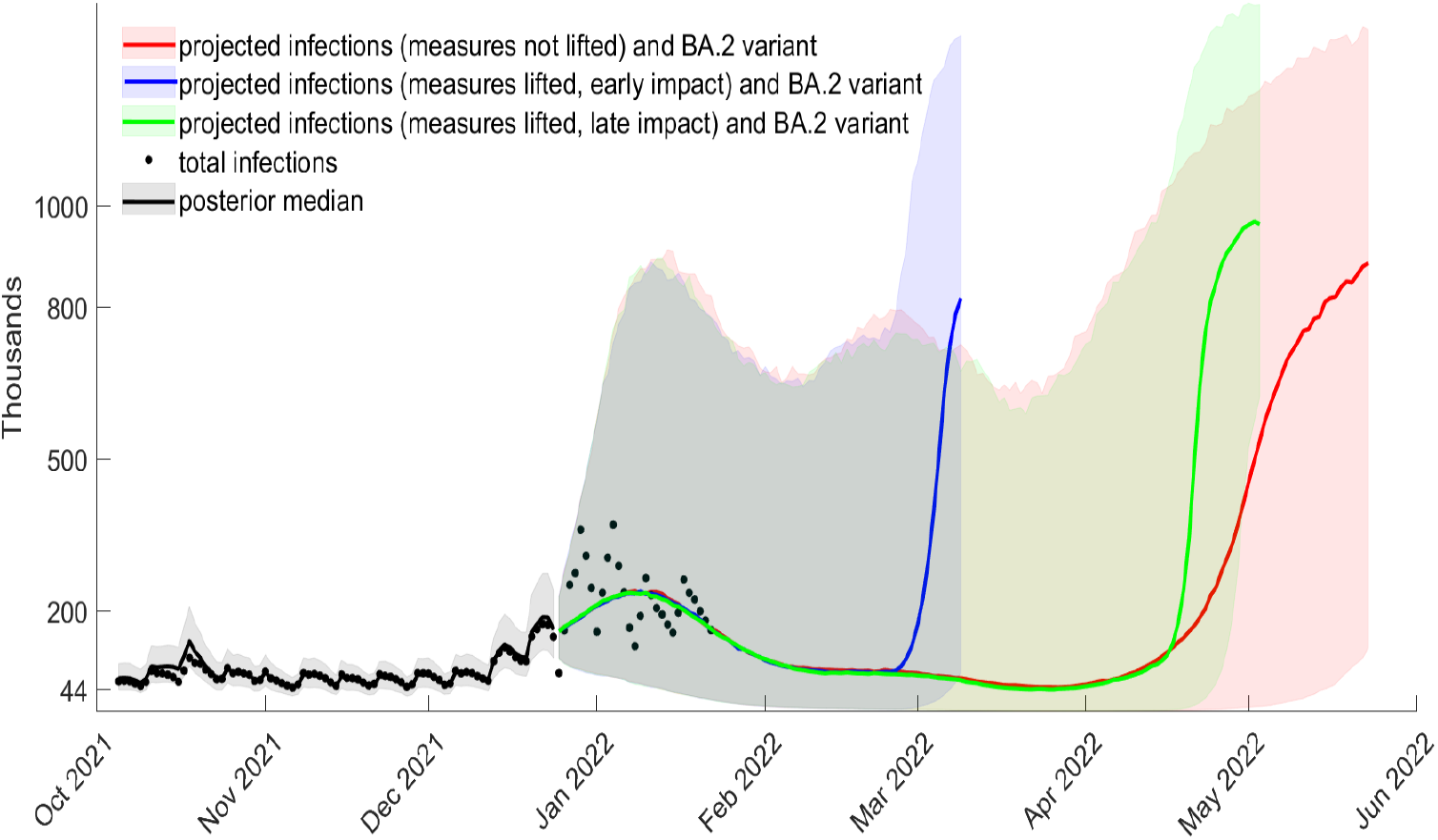
Projected total cases from December 25, 2021, waning of boosters after 6 months, *bir* = 0.75, *ρ*_*o*_ = 0.41 (posterior median), relative BA.2 intensity increase *ρ*_*BA*.2_ = 20%, with the lower 5% to the upper 95% quantiles

### S6. Further results on the impact on hospital admissions

Figure S27 shows the projected new admission into hospital from January 2, 2022 based on the median of projected infections from Figures S21-S23 when the waning of the vaccine booster is after 6 months.

**Figure S27:**
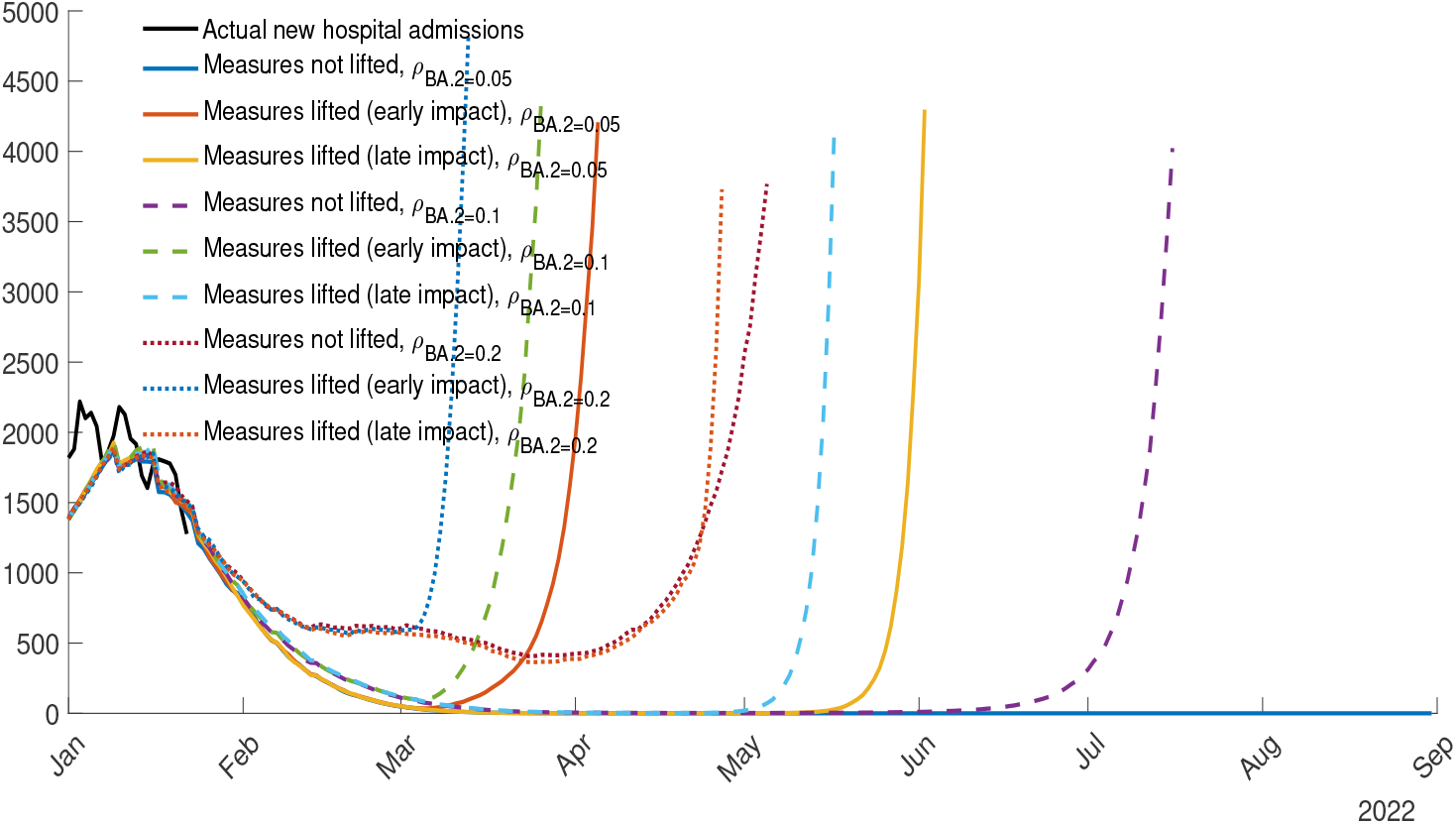
Projected new hospital admissions based on the median projected infections from Figures S21-S23, waning of boosters after 6 months, *bir* = 0.75, *ρ*_*o*_ = 0.41 (posterior median)

## Footnotes

1 See e.g. the Supplementary Material in Sonabend et al. (2021), which describes how they estimate an age-structured, regional, multiple vaccine type, multiple variant model.

2 We chose the negative binomial distribution as this is most commonly used to model overdispersion in epidemiology models (see, e.g., Rozhnova et al., 2021, Viana et al., 2021).

3 The dynamic INGARCH model was also used by Agosto and Giudici (2020), Roy and Karmakar (2021) and Giudici et al. (2021) to model COVID-19 infections in U.S and Italy, though without accounting for overdispersion. The first study assumes stationarity and constant parameters, therefore not accounting for NPIs. The second study models NPIs nonparametrically, with Bayesian B-Splines, which makes it difficult to establish which periods relate to a particular NPI. Giudici et al. (2021) use the Oxford COVID-19 Government Response Tracker to create NPI variables which are then included as exogenous variables in the model, but this approach ignores endogeneity of individuals’ responses to NPIs. Unlike our study, all three studies mentioned only use reported infections, and do not account for vaccination, waning of vaccines, or variants of concern.

4 To motivate this choice further, note that the probabilities *g*_*j,t*_ are unlikely to be identified within the dynamic intensity model, separately from the time-varying effect of vaccinations and NPIs. Additionally, Götz et al. (2021) show that if the number of susceptible individuals is fixed in an SIR (susceptible-infected-recovered) model with two virus strains, then *g*_*j,t*_ fitted to the share of the new variant in all cases within a period can be used to approximate the transition in infectiousness from the old variant to the new one. Hansen (2021) shows this as well, without using an SIR model. In both papers, the *κ*_*j*_ parameter directly relates to relative infectiousness of the new variant. We instead estimate *ρ*_*α*_, *ρ*_*δ*_ and *ρ*_*o*_ directly within the dynamic intensity model, following Viana et al. (2021) and therefore assuming that we reach an average new intensity when the transition is completed.

5 The first observation for the PCR test surveillance in random samples of the population is May 3, 2020.

6 The Supplementary Appendix, Section S1, shows the time-varying ratio of total to reported cases.

7 We stress here that *vir* and *bir* cannot be interpreted as vaccine and booster effectiveness against infection, as this is a term usually reserved for comparing vaccinated with non-vaccinated in a controlled setting.

8 Therefore, when *c >* 0.7, the vaccination speed is faster.

9 Plan B refers to measures introduced on December 8, 2021, in England: working from home for those who can, face mask wearing to most public indoor venues, vaccine passport, and daily tests for those who are contacts of Omicron cases.

